# Reward contamination in restrictive anorexia nervosa: A meta-analysis of functional MRI studies

**DOI:** 10.1101/2025.04.06.25325338

**Authors:** Charlotte S. Rye, Felippe E. Amorim, Laetitia H.E. Ward, Amy L. Milton

**Affiliations:** Department of Psychology, University of Cambridge, Downing Street, Cambridge, CB2 3EB; Department of Physiology, Development and Neuroscience, University of Cambridge, Downing Street, Cambridge, CB2 3EG

**Keywords:** anorexia, reward contamination, fMRI, anhedonia

## Abstract

Individuals with anorexia nervosa (AN) are typically anhedonic, leading to the suggestion that intrinsic disturbances of reward processing may represent a trait marker of the disorder. Previous studies have used task-based functional magnetic resonance imaging (fMRI) to investigate reward-related brain activity in AN and reported altered activation in the prefrontal cortex, dorsal posterior cingulate cortex, and rostral anterior cingulate cortex. However, likely due to the varied paradigms and methodologies used, as well as the heterogeneity in sample characteristics, results have proved inconsistent. To determine whether AN patients with the restrictive subtype (AN-r) show different reward-induced activation patterns to matched healthy controls (HCs) at different illness stages, we conducted a meta-analysis of 19 task-based fMRI studies of reward-processing. Using the seed-based differential mapping (SDM) technique, we found differences in reward-related brain activity between AN-r and HCs. Moreover, different brain regions showed differential activation across illness stages, with the direction and magnitude of effects dependent on specific task stimuli. These findings suggest that those with AN-r show distorted reward processing as a consequence of reward contamination and alterations in valence assignment to reward stimuli. In weight-recovered AN-r patients, differences to HCs persisted but were limited to regions known to exhibit significant atrophy in AN-r, indicating that altered reward processing is associated with anorectic undernutrition. These findings have implications for developing pharmacological treatments to aid psychological recovery in AN-r.

## 1. INTRODUCTION

Anorexia nervosa (AN) is an eating disorder characterised by a restriction of energy intake relative to requirements and an intense fear of gaining weight (American Psychiatric Association, 2013). Recent findings suggest that the incidence of AN is increasing in both males and females (van Eeden et al., 2021), yet prognosis remains poor (Bischoff-Grethe et al., 2013) and AN has the highest mortality rate among psychiatric disorders (Zipfel et al., 2015). There is therefore a large unmet clinical need for more effective and accessible evidence-based treatments.

AN can be classified into restricting (AN-r) and binge-purge (AN-bp) subtypes which have been reported to differ regarding their underlying pathophysiology (Van Autreve et al., 2013) and behavioural patterns (Murao et al., 2017).^1^ Whilst individuals with AN-r restrict their food intake and commonly engage in excessive physical activity, individuals with AN-bp regularly engage in binge eating and/or purging behaviours (Murao et al., 2017). Psychiatric comorbidities also differ between subtypes: individuals with AN-bp have higher rates of affective disorders like major depression (Herzog et al., 1992), whilst avoidant personality disorder is more commonly observed in AN-r (Diaz-Marsa et al., 2000).

Although the aetiology of AN remains unclear, previous behavioural (O’Hara et al., 2016) and neuroimaging (Fladung et al., 2013) studies have indicated that altered reward processing and cognitive control processes may underlie AN symptomatology, particularly AN-r (Wilson et al., 2019). Whilst it is important to acknowledge that factors beyond reward-related processes are implicated in eating disorders, a greater understanding of reward abnormalities in AN-r and their relevance to illness symptoms could facilitate the development of novel pharmacological interventions translated from basic neuroscience (Nord et al., 2021), making this an important research area.

One possibility is that abnormalities in brain reward function may be a causal factor in the complex aetiology of AN-r (Berridge, 2009). Alternatively, distortions in reward processing may be a secondary consequence of disordered intake, representing a state marker of acute undernutrition (King et al., 2020). Even if this is the case and distortions to reward processing are a consequence rather than a cause of AN-r, they may still provide a target for pharmacological treatments to normalise eating behaviour through modulation of reward function.

Individuals with AN-r are often considered anhedonic (Wagner et al., 2007), and typically describe primary rewards as unpleasant (Ehrlich et al., 2015). However, since they appear to value core symptoms such as hunger that many would find aversive (Sweitzer et al., 2018), this apparent anhedonia may instead be interpreted in line with Reward Contamination Theory (RCT). First proposed by Keating (2010), RCT attempts to explain the observation that many patients with AN experience normally rewarding stimuli (e.g., food) as punishing, and punishing stimuli (e.g., self-starvation) as rewarding. RCT posits that the behaviours engaged in by patients such as self-starvation and excessive exercise are initially rewarding since they allow for the maintenance of the desired low body weight but become reinforced in a pathological manner (Sodersten et al., 2016). However, patients fail to recognise that aspects of reward are being contaminated with punishment, resulting in abnormal attribution of stimulus valence and an inability to appropriately regulate their behaviours (Keating, 2010). In this manner, RCT posits that AN-r is not characterised by anhedonia, but rather alterations in the assignment of stimulus valence such that hunger loses its ability to increase the motivational drive for food consumption (Gizewski et al., 2010).

Whilst several functional MRI (fMRI) studies have used task-based paradigms to probe the functioning of the brain reward system in AN, the literature on altered reward processing in patients with AN-r remains somewhat contradictory, with the precise involvement of brain reward systems in AN-r remaining contested (Zhong et al., 2023).

Early studies found undifferentiated responses to monetary reward in the ventral striatum (Wagner et al., 2007), yet more recent studies have reported increased (Bischoff-Grethe et al., 2013) or even decreased (Sweitzer et al., 2018) activity in reward-related regions. Such heterogeneous results may in part result from differences in study design, patient inclusion/exclusion criteria and small sample sizes. For example, whilst the neurobiological hypothesis of AN posits that the clinical phenomena of AN can be accounted for by a dysfunctional insula restricting the integration of bodily and external signals (Nunn et al., 2008), this has only been validated in studies using food-based stimuli (e.g. Wagner et al., 2008). Meta-analytic tools, which serve as powerful instruments for analysing cumulative data from studies with limited sample sizes and low statistical power, therefore have great potential to further our understanding of AN-r pathology. To our knowledge, only two meta-analyses have investigated brain activation using tasked-based fMRI studies in patients with AN-r (Zhong et al., 2023; Zhu et al., 2012), and neither considered the effect of AN-r clinical staging (i.e., acute, chronic, recovered) on reward-based neural activation. This is an important issue since the course of AN-r onset, progression and recovery shows significant individual variation (Espel-Huynh et al., 2020), and a greater understanding of how AN progresses should guide future research and development of interventions. Furthermore, acute starvation can have significant effects on cognitive functioning, with Lozano-Serra and colleagues (2014) highlighting the importance of weight-related hormonal function in perceptual organisation, memory, information processing and cognitive flexibility. Thus, the metabolic effects of low food intake may produce artificial differences in acute and chronic patients compared to recovered individuals.

In their meta-analysis, Zhu et al. (2012) used an activation likelihood estimation (ALE) analysis which gives greater weight to studies with a higher number of neural foci and relies on spatial assumptions that may not be met by the included studies (Radua et al., 2012). To this end, we conducted a meta-analysis of neuroimaging studies using the seed-based differential mapping (SDM) technique. The SDM technique has been fully validated in several studies (e.g., Su et al., 2021) and has greater power than approaches such as ALE (Radua et al., 2012).

The overarching aims of this meta-analysis were to investigate (i) whether reward processing is abnormal in AN-r (and if so, whether alterations are more in keeping with a model of anhedonia or reward contamination) and; (ii) whether reward-associated brain activation differs as a function of illness stage (acute, chronic, recovered).

## 2. METHODS

The protocol related to this meta-analysis was registered prior to data-collection on the Open Science Framework (OSF) (https://doi.org/10.17605/OSF.IO/4JVZA). The meta-analysis adhered to the Preferred Reporting Items for Systematic Reviews and Meta-Analyses (PRISMA) checklist (Page et al., 2021).

### 2.1. Literature Search

Thorough searches of the Web of Science (1^st^ January 1994 – 31^st^ December 2023) and PubMed (1994-2023) were conducted, and the reference lists of any included studies were screened to identify any additional relevant studies. The following Medical Subject Heading (MeSH) terms were used for the search: (“anorexi*”) AND (“functional magnetic resonance imaging” OR “fMRI”) AND (“reward” OR “reinforce*”). For the initial screening, three independent investigators (C.S.R., F.E.A. and L.H.E.W) screened studies using information available in titles and abstracts.

### 2.2. Study Inclusion Criteria

The following inclusion criteria had to be met: (i) articles on patients diagnosed with AN-r according to the Diagnostic and Statistical Manual of Mental Disorders, Fifth Edition (DSM-5) (American Psychiatric Association, 2013) or Fourth Edition (DSM-IV) (American Psychiatric Association, 1994) criteria who had no co-occurring alcohol or drug misuse; (ii) articles involving reward-related, task-based fMRI studies with a healthy control (HC) comparison group; (iii) primary studies limited to human subjects reported in an English language peer-reviewed journal; (iv) articles reporting the peak coordinates of significant clusters in three-dimensional coordinates (Talairach or Montreal Neurological Institute [MNI]). Where studies reported results from both AN-r patients and those with AN-bp, only data comparing the AN-r and HC groups was considered. If studies reported data from both current and recovered AN-r, these were considered as separate datasets. Articles were excluded from the analysis if: (i) the publication included data that overlapped with another; (ii) there was no HC group; (iii) neuroanatomical coordinates or other data were unavailable even after contacting the authors (**Figure 1**). When discrepancies were detected, the final decision regarding study inclusion was made by a fourth investigator (A.L.M.). From an initially identified 206 English language, peer-reviewed articles, 15 primary studies, containing 18 datasets we included in the analysis (**Figure 1**). All included studies were approved by ethical committees at the respective institutions, and protocols complied with national legislation, with the privacy rights of all participants observed. All participants over the age of 18 gave written informed consent. Participants under the age of 18 gave written informed assent, and their parents gave written informed consent.

**Figure 1:**
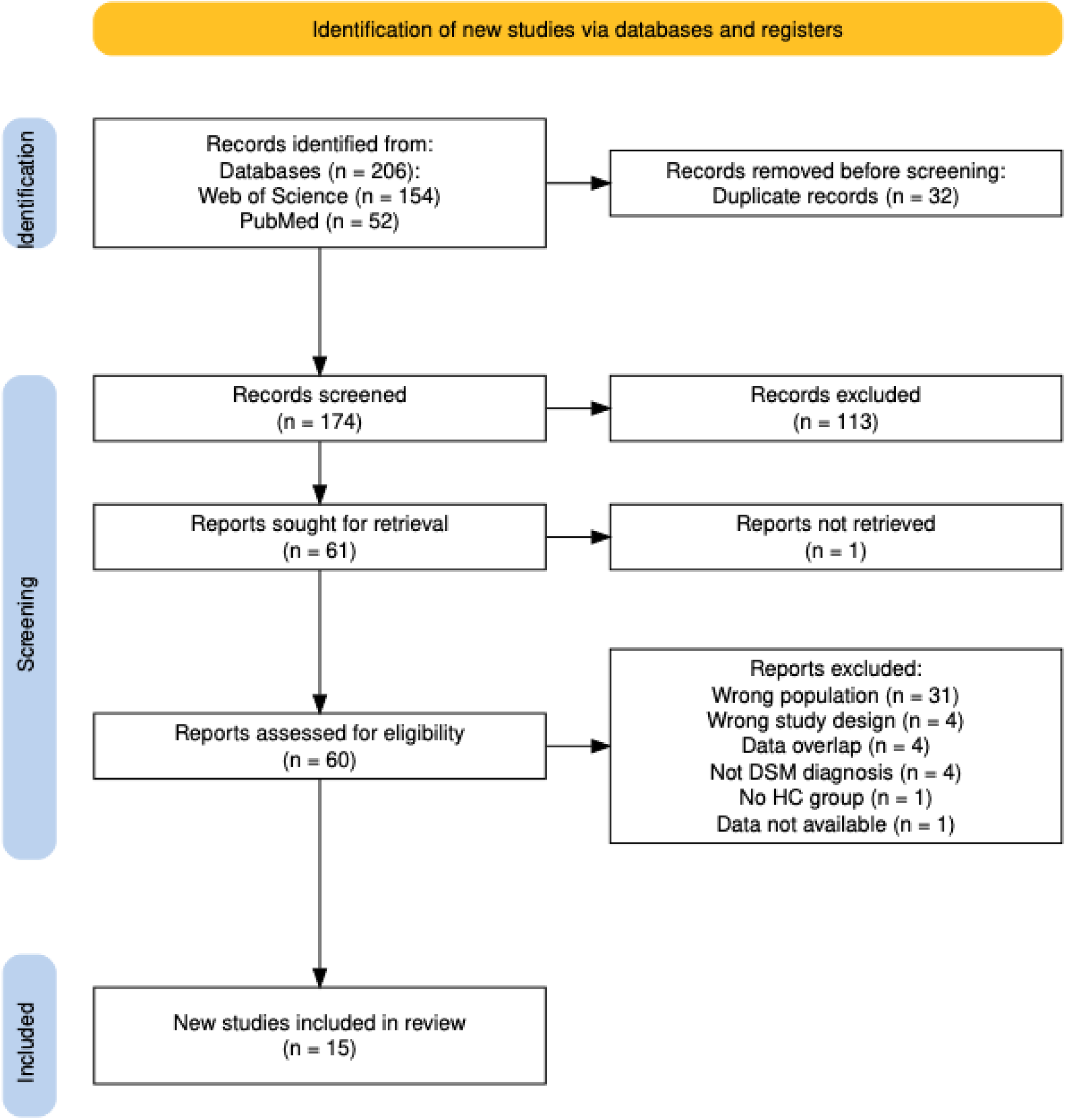
PRISMA flowchart of the literature search and study selection process. From 206 identified records, 15 reward-based fMRI studies of restrictive anorexia nervosa (AN-r) patients (containing 18 datasets) met the criteria for inclusion in the meta-analysis.

### 2.3. Quality Assessment

The following data was extracted from each included study: surname of the first author, year of publication, sample size (AN-r and HC), mean (±SD) age, mean (±SD) BMI, mean (±SD) illness duration, gender composition, any treatment, whether AN-r was current or recovered (if recovered mean (±SD) months since beginning of recovery), software package used for image processing, effect sizes (*g*), nature of the task completed, peak coordinates (MNI) and peak heights (*t*-value). The quality of each included study was assessed using the 12-point checklist developed by Zhong et al. (2023) (**Supplementary Material 1**).

### 2.4. Data Analysis

Meta-analysis was conducted using the SDM-PSI software package (version 6.22) which allows for meta-analysis of voxel-based neuroimaging studies and weighting of calculations for intra-study variance and inter-study heterogeneity. Using the SDM approach, study weights are proportional to the number of participants in the study, and effect sizes are used to combine reported peak coordinates that are extracted from the datasets (Radua et al., 2012). This independent consideration of effect sizes allows meta-regression analyses to be conducted.

SDM-PSI was used to incorporate all relevant information from the individual primary studies into a single map based on the reported peak coordinates and *t*-values. Analysis was performed as described by Radua et al. (2012). The default SDM kernel size and thresholds (full anisotropy = 1, isotropic full width at half maximum [FWHM] = 20mm, and voxel = 2mm) were used to ensure optimal balance of type I and type II errors (Radua et al., 2014). Results were reported using an uncorrected *p* = .005 threshold and minimum cluster extent = 50 voxels as this has been validated as a strong approximation of corrected results (*p* < .05) (Radua et al., 2012). The main meta-analysis was conducted using all identified datasets (n=18). Subgroup meta-analyses were also conducted, including (i) only those with current, acute AN-r (illness duration <60 months; n = 3), (ii) only those with current, chronic AN-r (mean illness duration >60 months; n = 5), (iii) only those who had recovered from AN-r (patients who had maintained a healthy BMI and had not engaged in restrictive eating patterns for at least 6 months prior to the study; n=9). The cut-off of 60 months illness duration was chosen to define the acute and chronic groups as this is the mode in the literature (Broomfield et al., 2017). Due to the limited number of datasets available, it was not possible to conduct further subgroup analyses with both task stimulus and illness stage as factors.

AN-r is highly comorbid with anxiety and mood disorders, both of which have been associated with dysfunctional reward processing (Dillon et al., 2014). Moreover, treatment with widely prescribed psychiatric medications such as selective serotonin reuptake inhibitors (SSRIs) can alter the neural processing of both rewarding and aversive stimuli (McCabe et al., 2010). Therefore, exploratory analyses were conducted (i) excluding studies reporting comorbidities (n=8), (ii) excluding studies reporting the use of psychotropic medications (n=8), (iii) taking task stimulus as a covariate (food-related, n=11; monetary reward, n=6; social judgement, n=1).

#### 2.4.1. Analysis of Heterogeneity and Publication Bias

A heterogeneity analysis was conducted, with I^2^ statistics used to assess between-study robustness. A value of I^2^ < 50% indicates low heterogeneity (Egger, Smith, & Phillips, 1997). Egger’s tests were performed to examine publication bias (Egger et al., 1997). This involves quantitative analysis by extracting the values from significant peaks, with the significance level set to *p* < .05.

#### 2.4.2. Meta-Regression Analysis

Meta-regression analyses were carried out assuming a mixed-effects model to explore the associations between the analytic results and clinical characteristics including age, illness duration, age at onset (AAO) and year of publication. To minimise the reporting of spurious findings, the *p*-value threshold was set to .0005.

## 3. RESULTS

### 3.1. General Overview

A total of 15 reward-related fMRI studies with primary data available were identified (**Figure 1**). Three included both active and recovered AN-r patients, leading to a total of 18 independent samples for inclusion in the meta-analysis. The identified studies included 339 AN-r patients (all females; mean age 24.8±4.6 years; mean illness duration 70.2 months; mean BMI 18.0±2.9) and 307 HCs (all females; mean age 24.2±4.5 years; mean BMI 21.6±0.7). There was no significant difference in age between participants with AN-r and HCs (t_(31)_ = 0.43, p = .67). However, as predicted, a Mann-Whitney U test revealed a significant difference in BMI between individuals with (or recovered from) AN-r (*x̃* = 17.4) and controls (*x̃* = 21.5) (U = 13.5, p < .0001). A summary of clinical and imaging characteristics is shown in **Table 1**.

**Table 1:** Demographic, clinical and imaging characteristics of the included studies. Abbreviations: AN-r = anorexia nervosa patients; HCs = healthy controls; BMI = body mass index; NA = not available; SPM = statistical parametric mapping; MRI = Magnetic Resonance Imaging; FSL = FMRIB’s Software Library, the University of Oxford; AFNI = Analysis of Functional NeuroImages; JMP = “John’s Macintosh Project”, SAS Institute; FWE = family wise error; FDR = false discovery rate.

| Study | n AN-r | n HC | Mean ± SD<br>Age (years)<br>AN-r | Mean ± SD Age<br>(years) HC | Mean ± SD<br>BMI AN-r | Mean ± SD<br>BMI HC | Mean ± SD<br>Illness Duration<br>(months) | Mean ± SD Age<br>at Onset | Recovered (if yes<br>mean ± SD recovery<br>duration (months)) | Task | Comorbidities | Medication | Scanner | Software | Threshold | Quality<br>Score |
| --- | --- | --- | --- | --- | --- | --- | --- | --- | --- | --- | --- | --- | --- | --- | --- | --- |
| Frank et al.<br>(2016) | 21 | 27 | 22.9±6.1 | 26.2±7.0 | 16.0±1.1 | 21.5±1.4 | NA | NA | N | Food Task - taste | Yes | Yes | 3.0T MRI | SPM5 | Pfwe < 0.05 |  |
| Jiang et al.<br>(2019) | 14 | 12 | 24.9±4.7 | 24.1±3.1 | 15.8±1.0 | 21.5±2.7 | NA | NA | N | Food Task - Odours | No | No | 1.5T MRI | SPM5 | Puncorrected < .001 | 9.5 |
| **Acute** |  |  |  |  |  |  |  |  |  |  |  |  |  |  |  |  |
| Bischoff-Grethe<br>et al. (2013) | 10 | 12 | 16.2±1.8 | 15.4±1.6 | 16.4±1.4 | 21.5±1.7 | 31.1±26.1 | NA | N | Monetary Reward | NA | Yes | 3.0T MRI | AFNI | Puncorrected < .05 | 10 |
| Geisler et al.<br>(2017) | 36 | 36 | 16.0±2.6 | 16.3±2.6 | 14.7±1.3 | 20.4±2.5 | Acute (<36) | NA | N | Monetary Reward –<br>probabilistic reversal<br>learning | Yes | No | 3.0T MRI | SPM8 | Pfwe < .001 | 11.5 |
| Holsen et al.<br>(2012) | 12 | 11 | 21.8±2.7 | 21.6±1.3 | 18.0±0.8 | 22.4±1.3 | 60.0±32.4 | 16.7±3.1 | N | Food Task – visual<br>stimuli | Yes | Yes | 3.0T MRI | SPM8 | Pfwe < .05 | 10.5 |
| **Chronic** |  |  |  |  |  |  |  |  |  |  |  |  |  |  |  |  |
| Frank et al.<br>(2012) | 21 | 23 | 22.5±5.8 | 24.8±5.6 | 16.1±1.1 | 21.5±1.4 | 77.8±63.5 | NA | N | Food Task –<br>prediction error | Yes | No | 3.0T MRI | SPM5 | Pfwe < .05 | 11.5 |
| McFadden et al.<br>(2014) | 20 | 24 | 22.9±5.7 | 27.4±6.3 | 16.0±1.1 | 21.6±1.7 | 81.1±74.8 | 16.15±2.56 | N | Taste Conditioning | Yes | Yes | 3.0T MRI | SPM8 | Pfdr < .01 | 10.5 |
| Murao et al.<br>(2017) | 11 | 20 | 30.9±10.1 | 33.2±8.8 | 12.0±1.3 | 21.4±3.4 | 140.4±84.0 | NA | N | Monetary Reward –<br>incentive delay task | No | Yes | 3.0T MRI | SPM8 | Puncorrected < .001 | 11 |
| Scaife et al.<br>(2016) | 12 | 16 | 29.5±6.0 | 24.3±5.7 | 15.4±1.9 | 21.2±2.0 | 123.6±62.4 | 20.1±5.9 | N | Food Task – visual<br>stimuli | Yes | Yes | 3.0T MRI | FSL | Pfdr < .05 | 11.5 |
| Via et al. (2015) | 20 | 20 | 28.4±9.3 | 28.2±8.6 | 16.9±1.3 | 21.0±1.8 | 85.2±76.9 | 21.3±9.3 | N | Social Judgement | No | Yes | 1.5T MRI | SPM8 | Pfwe < .05 | 12 |
| **Recovered** |  |  |  |  |  |  |  |  |  |  |  |  |  |  |  |  |
| Cowdrey et al.<br>(2011) | 15 | 16 | 32.3±3.5 | 24.1±2.9 | 21.3±2.2 | 21.2±1.6 | NA | 14.7±1.9 | Y(42.0±28.6) | Food Task – taste | Yes | No | 1.5T MRI | SPM8 | Pfwe < .05 | 11.5 |
| Ehrlich et al.<br>(2015) | 30 | 30 | 22.0±3.2 | 21.6±3.2 | 21.2±1.9 | 21.3±2.2 | NA | 14.5±2.0 | Y (54.8±37.8) | Monetary Reward –<br>incentive delay task | No | No | 3.0T MRI | SPM5 | Puncorrected < .001 | 11 |
| Frank et al.<br>(2016) | 19 | 27 | 27.0±5.3 | 26.2±7.0 | 20.2±1.1 | 21.5±1.4 | NA | NA | Y (NA) | Food Task - taste | Yes | Yes | 3.0T MRI | SPM5 | Pfwe < 0.05 | 11 |
| McFadden et al.<br>(2014) | 24 | 24 | 30.3±8.1 | 27.4±6.3 | 20.8±2.4 | 21.6±1.7 | 70.8±62.5 | 16.6±2.4 | Y(94.8±72.1) | Taste Conditioning | Yes | Yes | 3.0T MRI | SPM8 | Pfdr < .01 | 10.5 |
| Ritschel et al.<br>(2017) | 31 | 31 | 22.3±2.8 | 22.1±3.0 | 21.0±1.9 | 21.3±2.1 | NA | NA | Y(NA) | Monetary Reward –<br>probabilistic reversal<br>learning | Yes | No | 3.0T MRI | SPM8 | Pfwe < .01 | 10.5 |
| Scaife et al.<br>(2016) | 14 | 16 | 24.3±5.7 | 24.3±5.7 | 20.9±1.6 | 21.2±2.0 | 69.6±50.8 | 16.5±2.1 | Y(NA) | Food Task – visual<br>stimuli | No | Yes | 3.0T MRI | FSL | Pfdr < .05 | 11.5 |
| Wagner et al.<br>(2007) | 13 | 13 | 26.6±6.8 | 26.4±6.9 | 20.7±2.4 | 22.6±2.0 | NA | NA | Y(NA) | Monetary Reward | No | No | 3.0T MRI | AFNI | Pcorrected < .05 | 10 |
| Wagner et al.<br>(2008) | 16 | 16 | 26.4±6.2 | 26.8±6.3 | 20.7±2.2 | 22.9±2.2 | NA | 16.4±3.1 | Y(45.7±30.2) | Food Task -<br>consumption | No | No | 3.0T MRI | JMP | Pfwe < .05 | 11 |

### 3.2. Pooled Meta-Analysis

Overall, there was a difference in reward-related brain activity between AN-r and HCs at the uncorrected *p* = .005 threshold and minimum cluster extent = 50. AN-r patients showed hyperactivity in right anterior thalamic projections (*g* = 0.19±0.009), left median cingulate/paracingulate gyri (*g* = 0.13±0.007), and the left middle frontal gyrus (*g* = 0.13±0.006) compared to HCs. In contrast, hypoactivation was observed in the left postcentral gyrus (*g* = −0.09±0.006), right hippocampus (*g* = −0.08±0.008), and right precuneus (*g* = −0.07±0.006), in AN-r patients compared to HCs (**Table 2**; **Figure 2**).

**Table 2:**
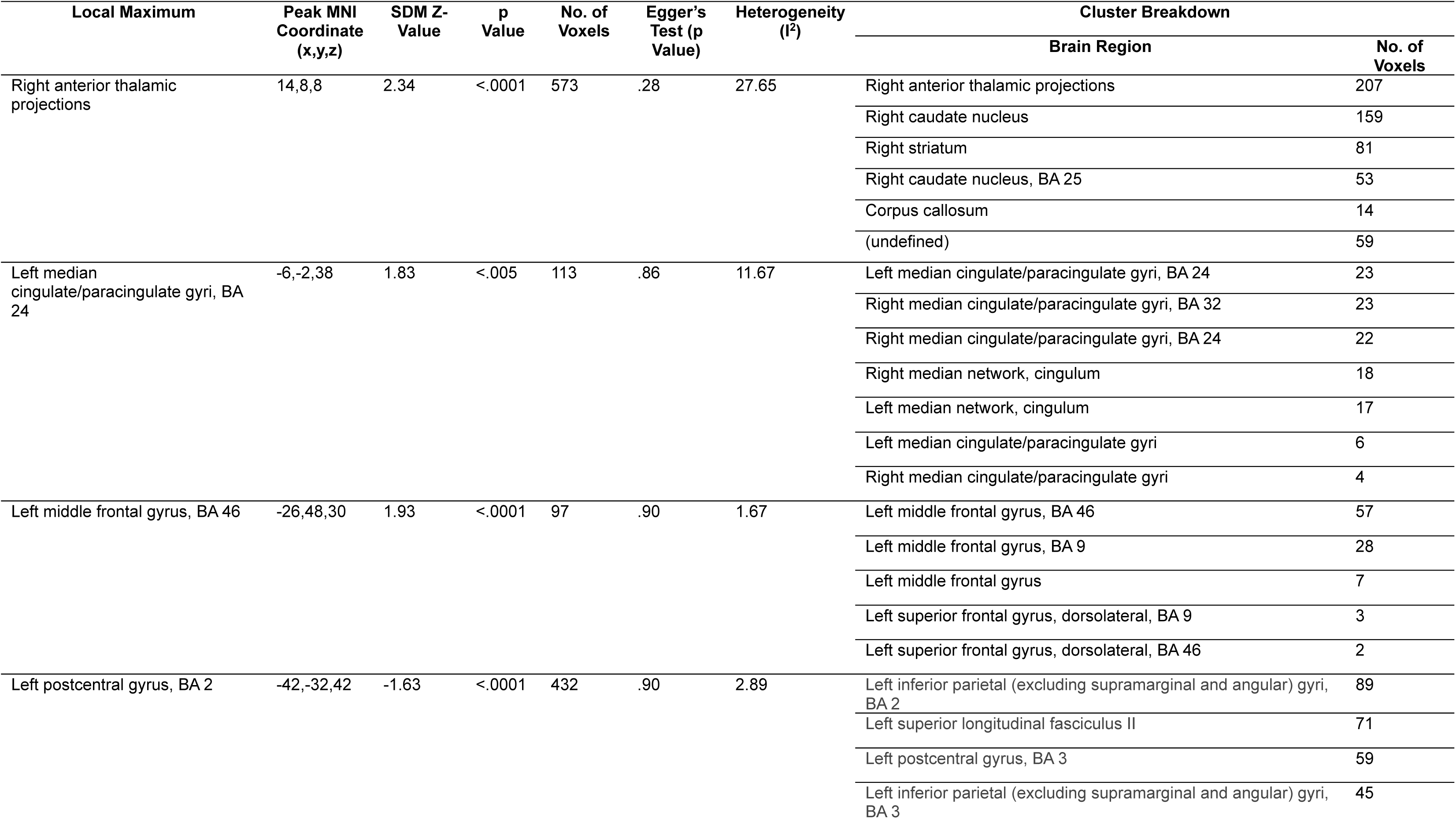
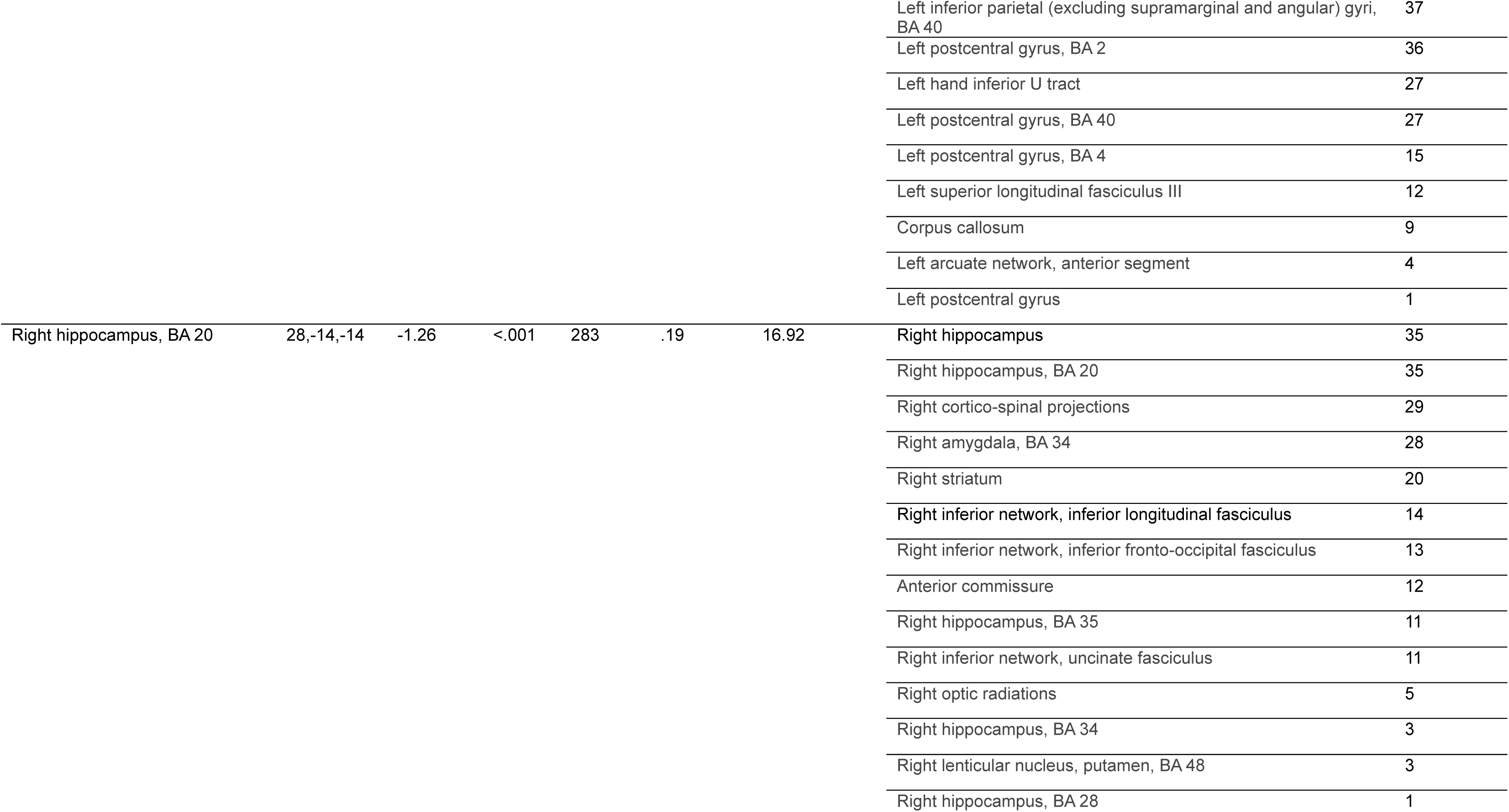
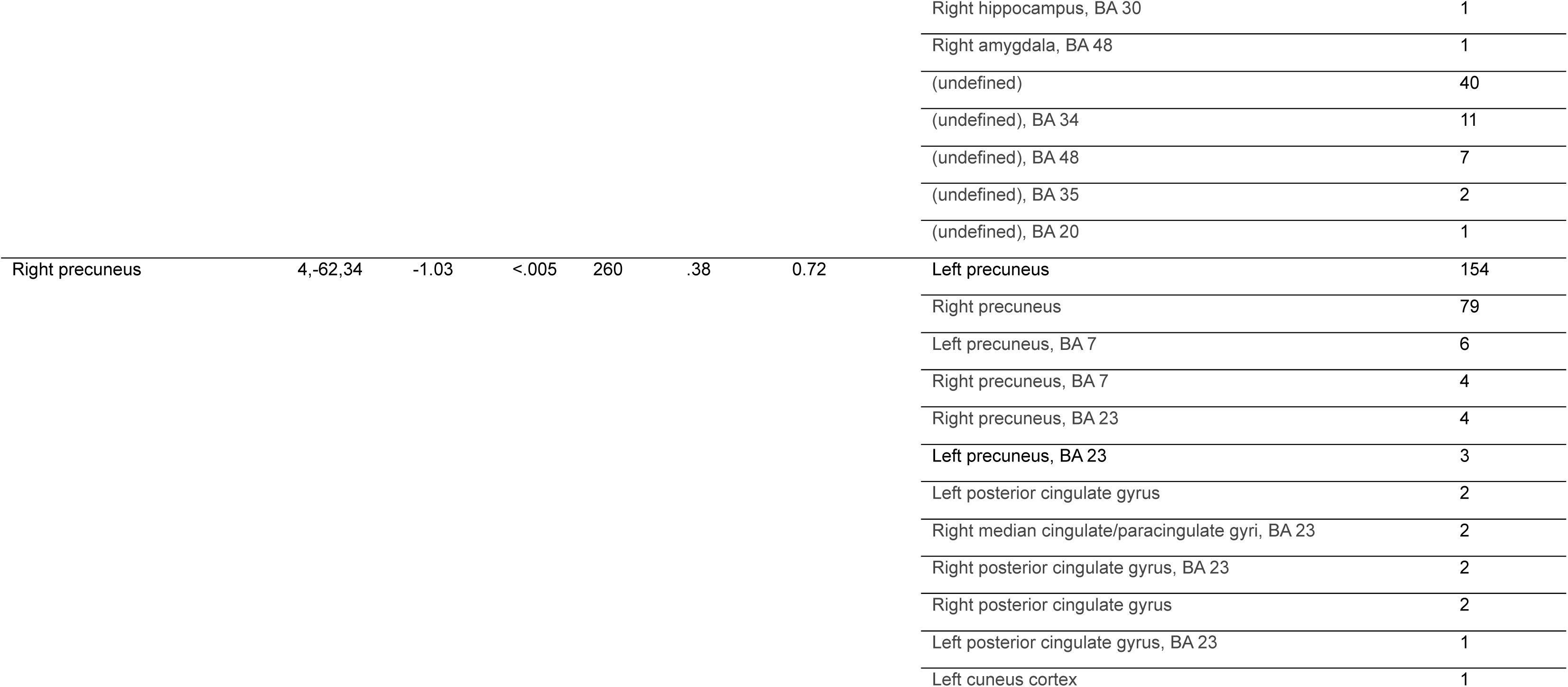
Meta-analysis of fMRI activity differences between individuals with AN-r and HCs. Abbreviations: fMRI = functional magnetic imaging; HCs = healthy controls; AN-r = restrictive anorexia nervosa; SDM = seed-based differential mapping; MNI =Montreal Neurological Institute; BA =Brodmann area.

**Figure 2:**
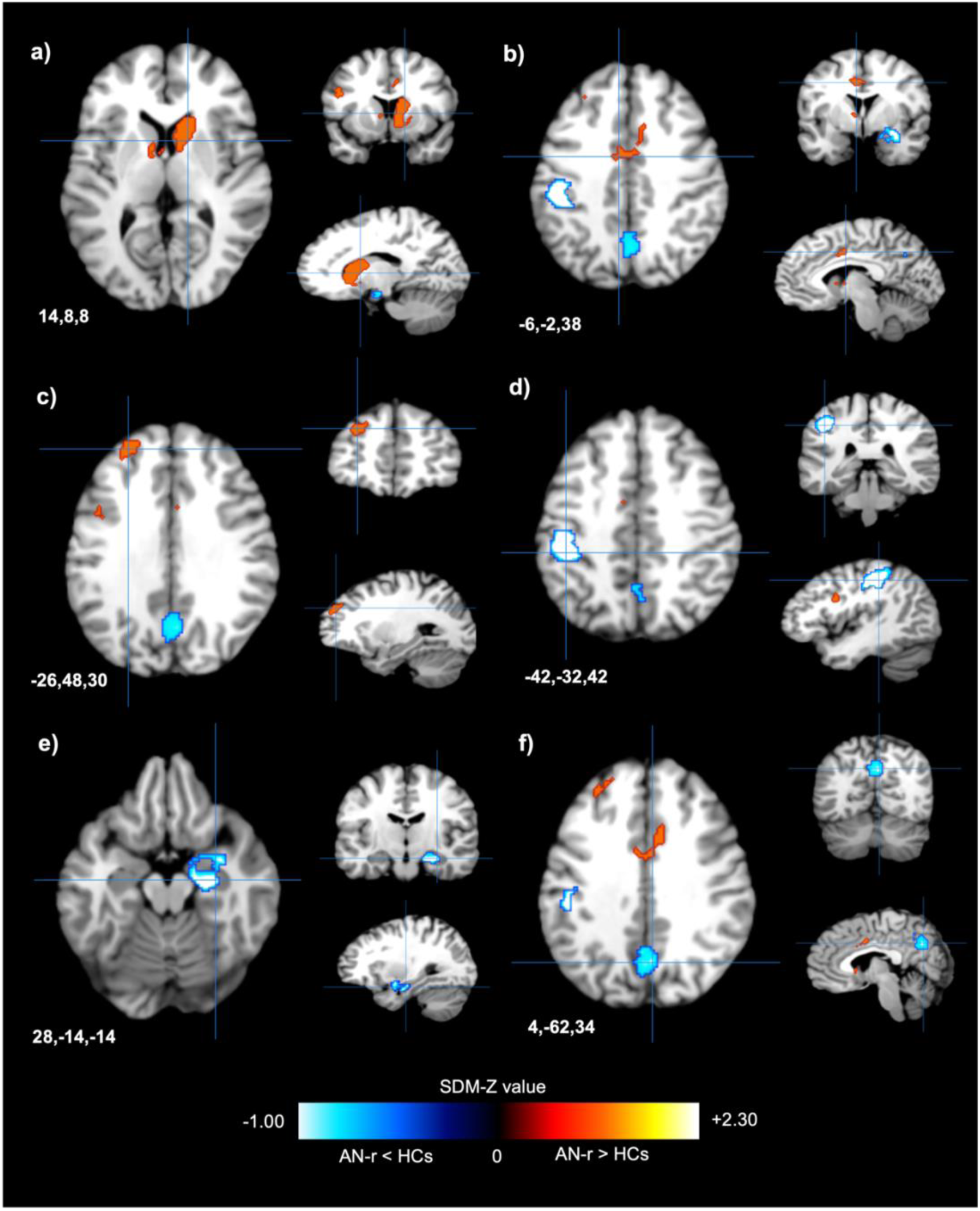
Patients with AN-r showed altered activity in reward-related brain regions as compared to HCs. **a.** Patients with AN-r showed greater functional activity in the right anterior thalamic projections (peak MNI coordinates x=14, y=8, z=8; max SDM-Z = 2.34) than HCs. **b.** Increased functional activity was observed in the left median cingulate/paracingulate gyri in AN-r (peak MNI coordinates x=-6, y=-2, z=38; max SDM-Z = 1.83). **c.** Patients with AN-r showed greater functional activity in the left middle frontal gyrus (peak MNI coordinates x=-26, y=48, z=30; max SDM-Z = 1.93) than HCs. **d.** Patients with AN-r exhibited hypoactivity in the left postcentral gyrus (peak MNI coordinates x=-42, y=-32, z=42; max SDM-Z = −1.63) compared to HCs. **e.** Patients with AN-r showed lower functional activity in the right hippocampus (peak MNI coordinates x=24, y=-14, z=-14; max SDM-Z = - 1.26) than HCs. **f.** An area of the right precuneus (peak MNI coordinates x=4, y=-62, z=34) showed decreased fMRI activity in AN-r (max SDM-Z = −1.03). Abbreviations: HCs = healthy controls; AN-r = restrictive anorexia nervosa patients; SDM = seed-based differential mapping; fMRI = functional magnetic resonance imaging.

### 3.3. Subgroup Meta-Analysis

Due to the effects of acute starvation on cognitive functioning (Green et al., 1996; Lozano-Serra et al., 2014), subgroup meta-analyses were conducted with illness stage as a factor. Two studies (Frank et al., 2016; Jiang et al., 2019) did not report illness duration for the active AN-r group and were consequently not included in subgroup analyses. Overall, there was no significant difference in effect size between subgroups (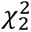^2^ = 0.81, df = 2, p = .67) (**Figure 3**). However, the brain regions showing altered activity did differ between subgroups.

**Figure 3:**
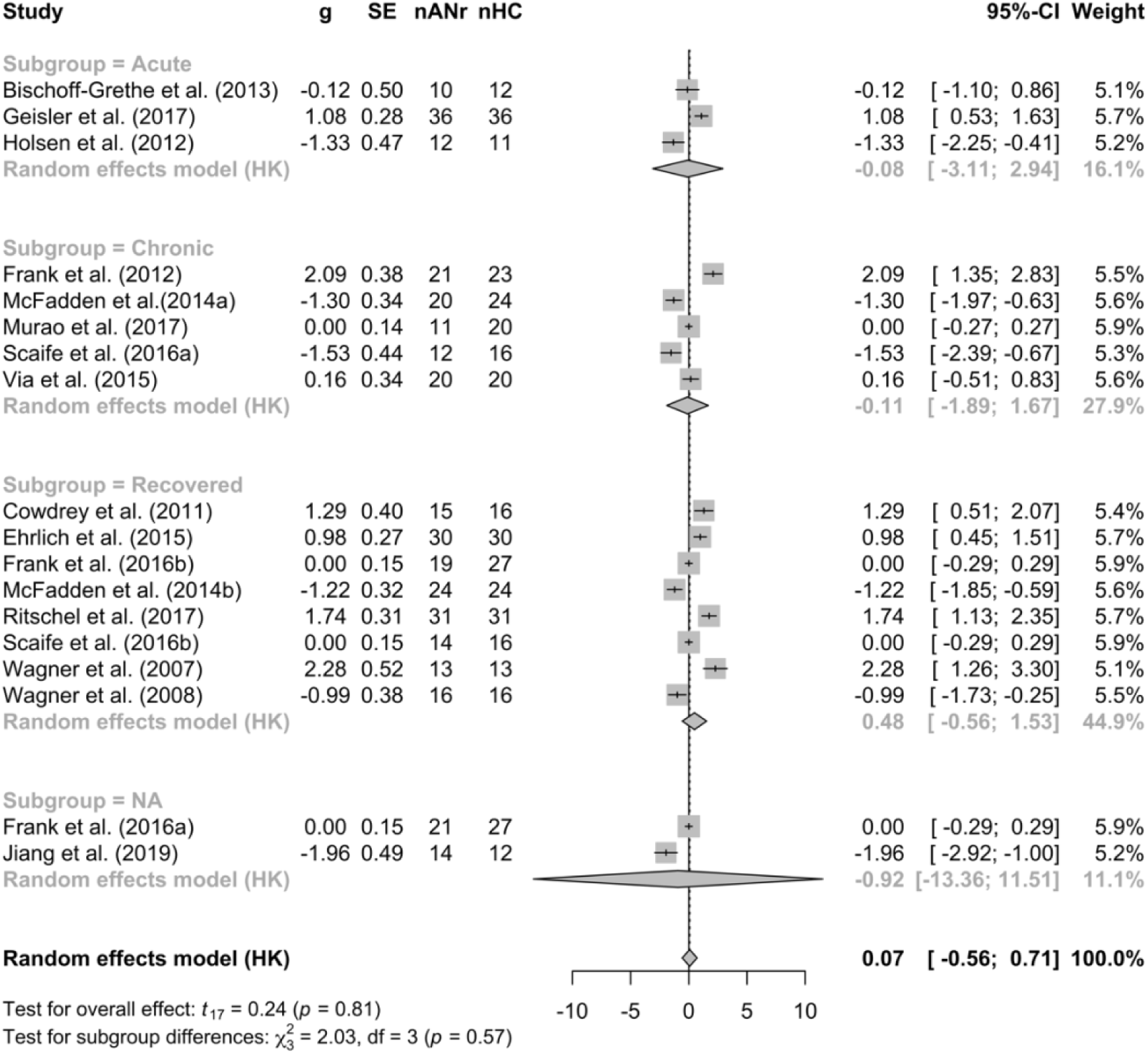
Forest plot showing the results of the pooled and subgroup random effects meta-analysis. Effect sizes (g) and standard error of effect sizes (SE) are shown for each included study and estimated effect size (g) and 95% confidence interval are given for each subgroup analysis and the pooled analysis. There was no significant overall effect and no difference in effect size between subgroups. Abbreviations: HCs = healthy controls; AN-r = restrictive anorexia nervosa.

#### 3.3.1. Acute Anorexia Nervosa

Analysis of studies of acute AN-r (<60 month illness duration) revealed that patients exhibited increased activity in the right median cingulate/paracingulate gyri (*g* = 0.46±0.05) and right anterior thalamic projections (*g* = 0.43±0.04) compared to HCs during reward processing tasks. Patients with acute AN-r also exhibited greater activation in the left middle frontal gyrus (*g* = 0.44±0.05) and right middle frontal gyrus (*g* = 0.39±0.04) compared to HCs. There were also regions of hypoactivity in AN-r patients during performance of reward-based tasks. These included the right amygdala (*g* = −0.37±0.06), left insula (*g* = −0.52±0.13) and left rolandic operculum (*g* = −0.40±0.08) (**Table 3**; **Figure 4**).

**Table 3:**
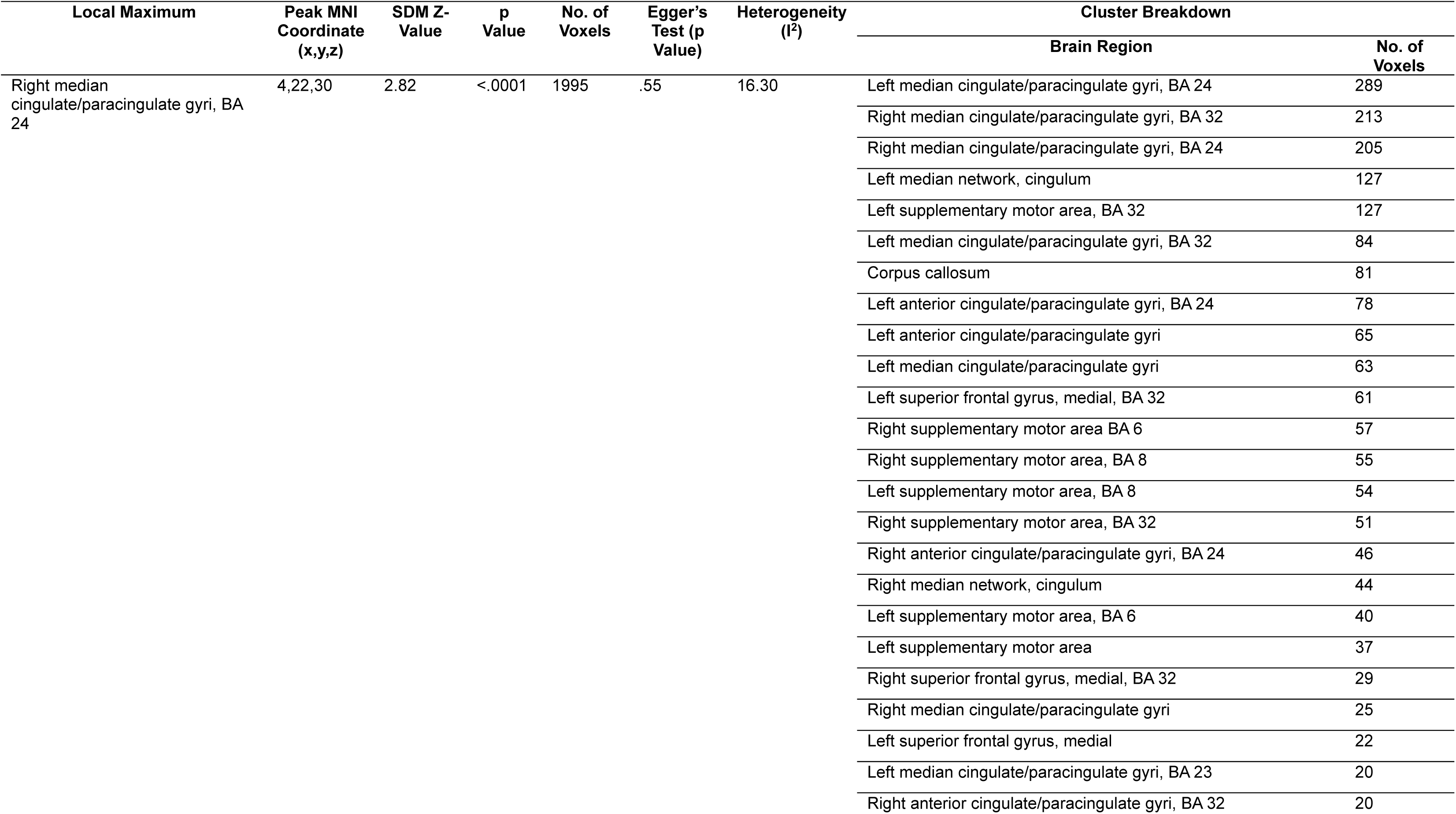
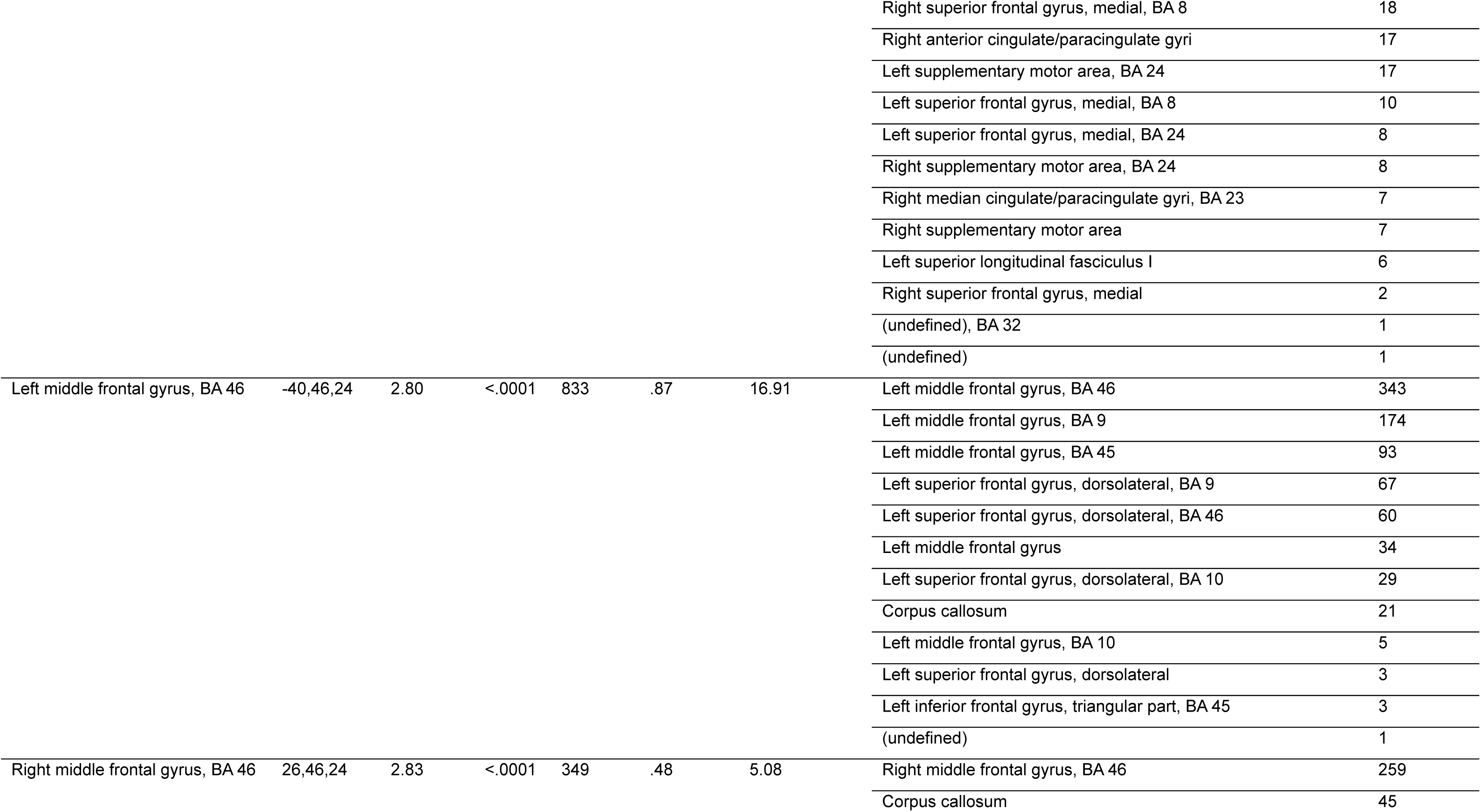
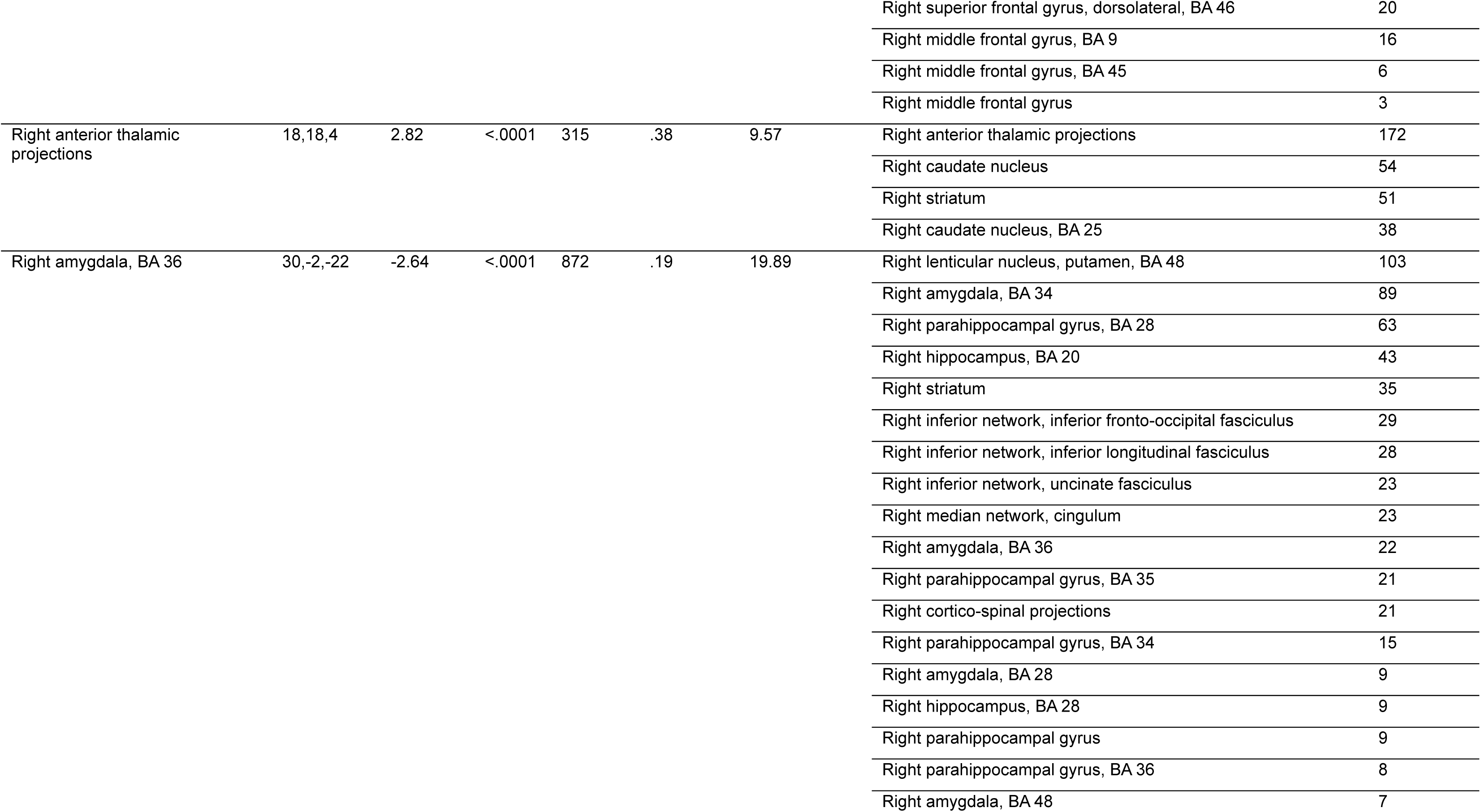
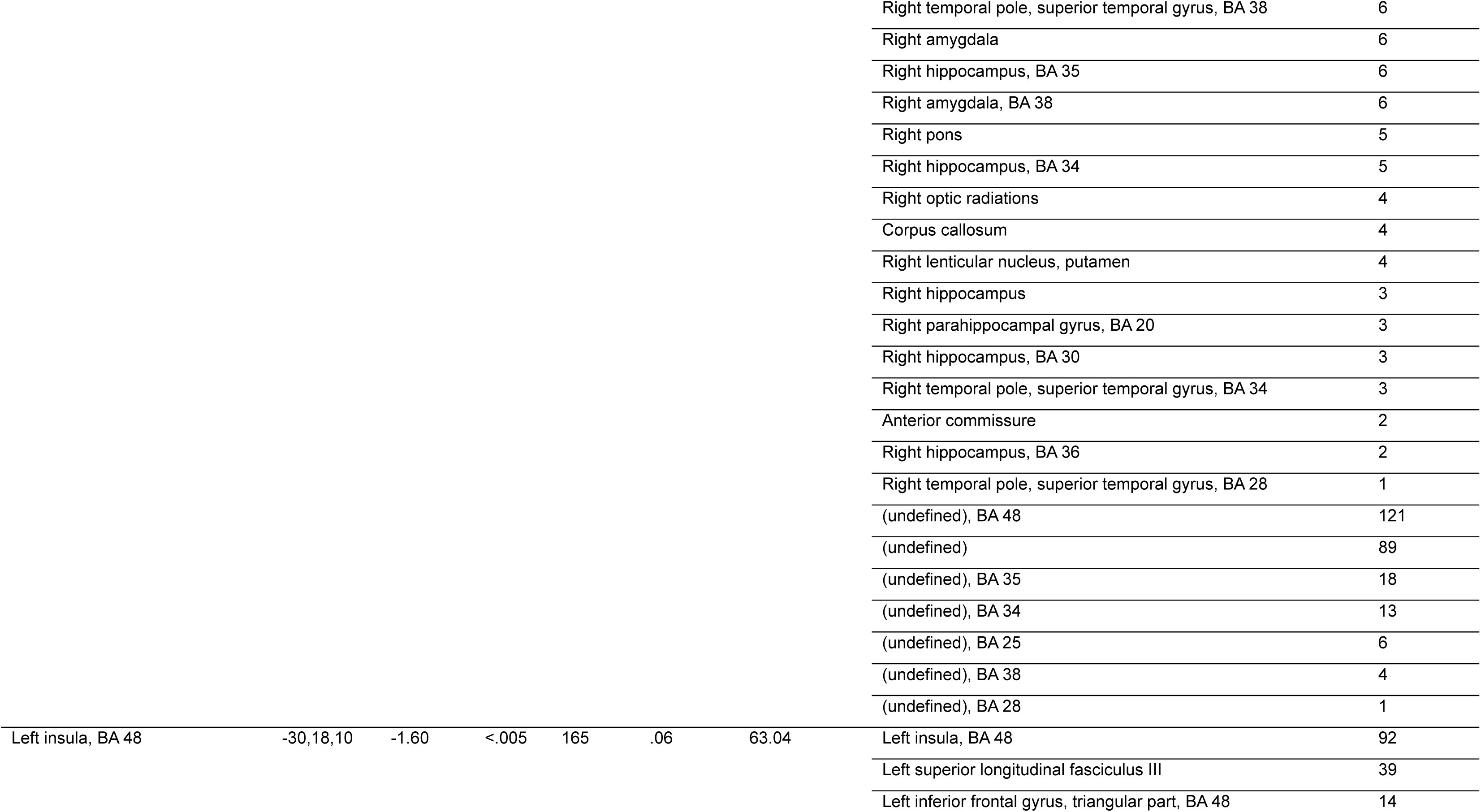
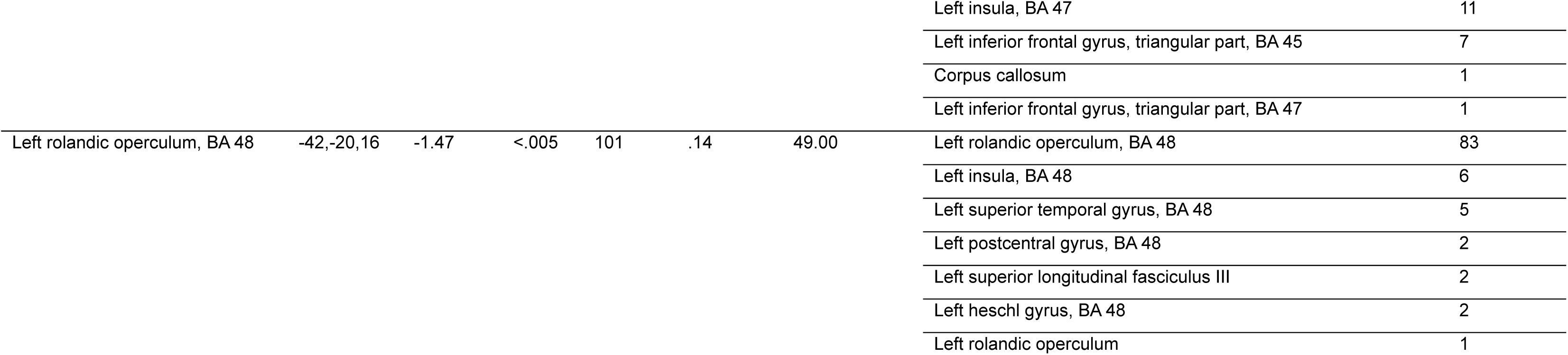
Meta-analysis of fMRI activity differences between individuals with current, acute AN-r and HCs. Abbreviations: fMRI = functional magnetic imaging; HCs = healthy controls; AN-r = restrictive anorexia nervosa; SDM = seed-based differential mapping; MNI =Montreal Neurological Institute; BA =Brodmann area.

**Figure 4:**
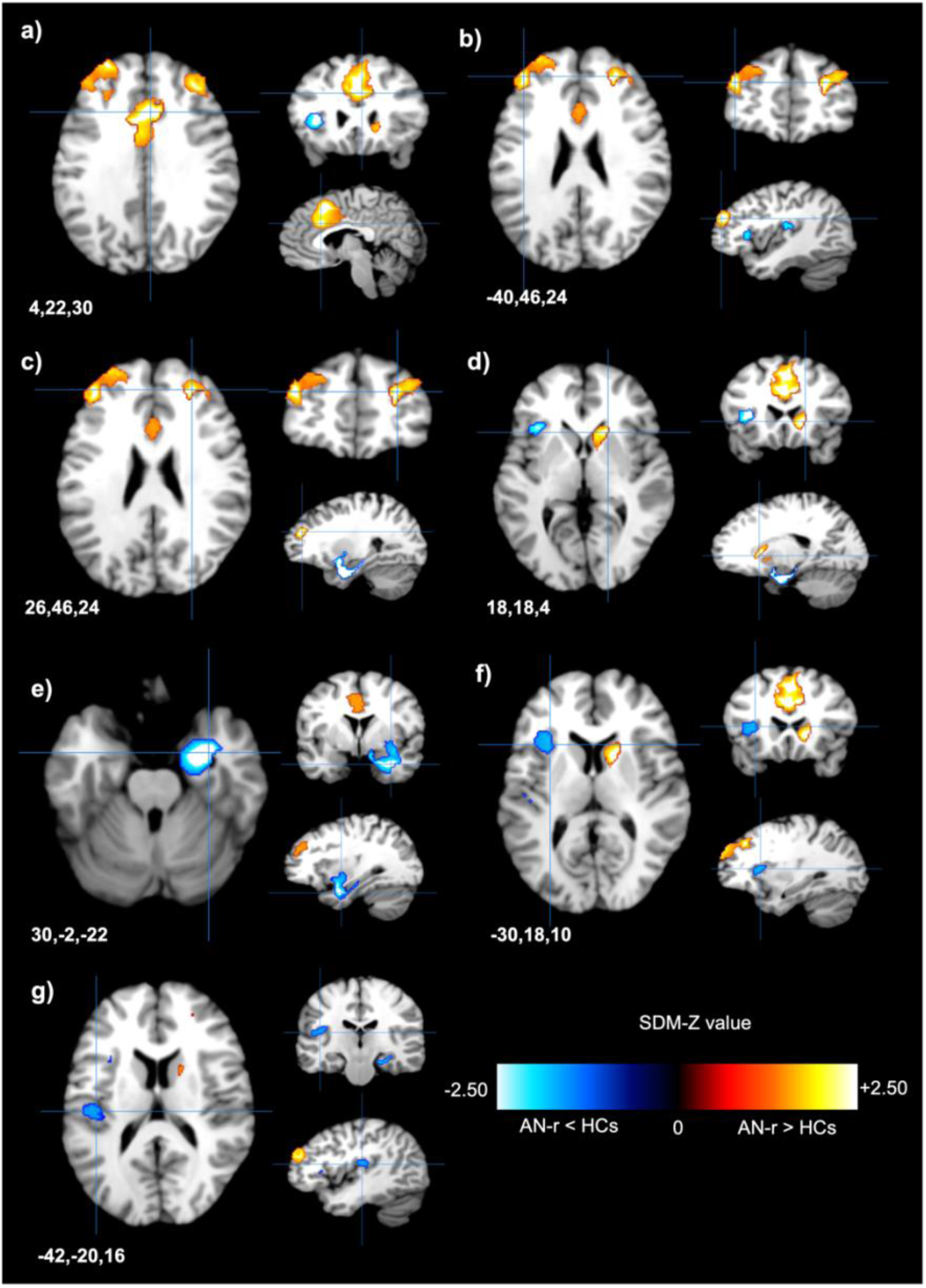
Patients with current, acute AN-r showed altered neural activity during reward-based tasks compared to HCs. **a.** Patients with acute AN-r showed greater functional activity in the right median cingulate/paracingulate gyri (peak MNI coordinates x=-4, y=22, z=30; max SDM-Z = 2.82) than HCs. **b.** Hyperactivity was observed in the left middle frontal gyrus (peak MNI coordinates x=-40, y=46, z=24; max SDM-Z = 2.80) in patients with acute AN-r compared to HCs. **c.** Patients with acute AN-r exhibited hyperactivity in the right middle frontal gyrus (peak MNI coordinates x=26, y=46, z=24; max SDM-Z = 2.83) compared to HCs. **d.** An area of the right anterior thalamic projections (peak MNI coordinates x=18, y=18, z=4) showed increased fMRI activity in acute AN-r (max SDM-Z = 2.82). **e.** Hypoactivity was observed in the right amygdala (peak MNI coordinates x=30, y=-2, z=-22; max SDM-Z = −2.64) in patients with acute AN-r compared to HCs. **f.** Hypoactivity was observed in the left insula (peak MNI coordinates x=-30, y=18, z=10; max SDM-Z = −1.60) in patients with acute AN-r compared to HCs. **g.** Patients with acute AN-r showed decreased functional activity in the left rolandic operculum (peak MNI coordinates x=-42, y=-20, z=16; max SDM-Z = −1.47) compared to HCs. Abbreviations: HCs = healthy controls; AN-r = restrictive anorexia nervosa patients; SDM = seed-based differential mapping; fMRI = functional magnetic resonance imaging.

#### 3.3.2. Chronic Anorexia Nervosa

Analysis of studies including only patients with current, chronic AN-r (>60 month illness duration) revealed a key region in the left middle occipital gyrus (peak MNI coordinate −20,-100,8) associated with significantly higher functional activity in patients than HCs (*g* = 0.29±0.04). Patients also exhibited higher activity in the right superior frontal gyrus (BA 10, peak at 8,60,0). However, this region did not exceed the minimum cluster extent to be considered significant. There were also significant differences in reward-related neural activity in the left postcentral gyrus (*g* = −0.20±0.03), with chronic AN-r patients exhibiting hypoactivity in this region compared to HCs (**Table 4**; **Figure 5**).

**Table 4:**
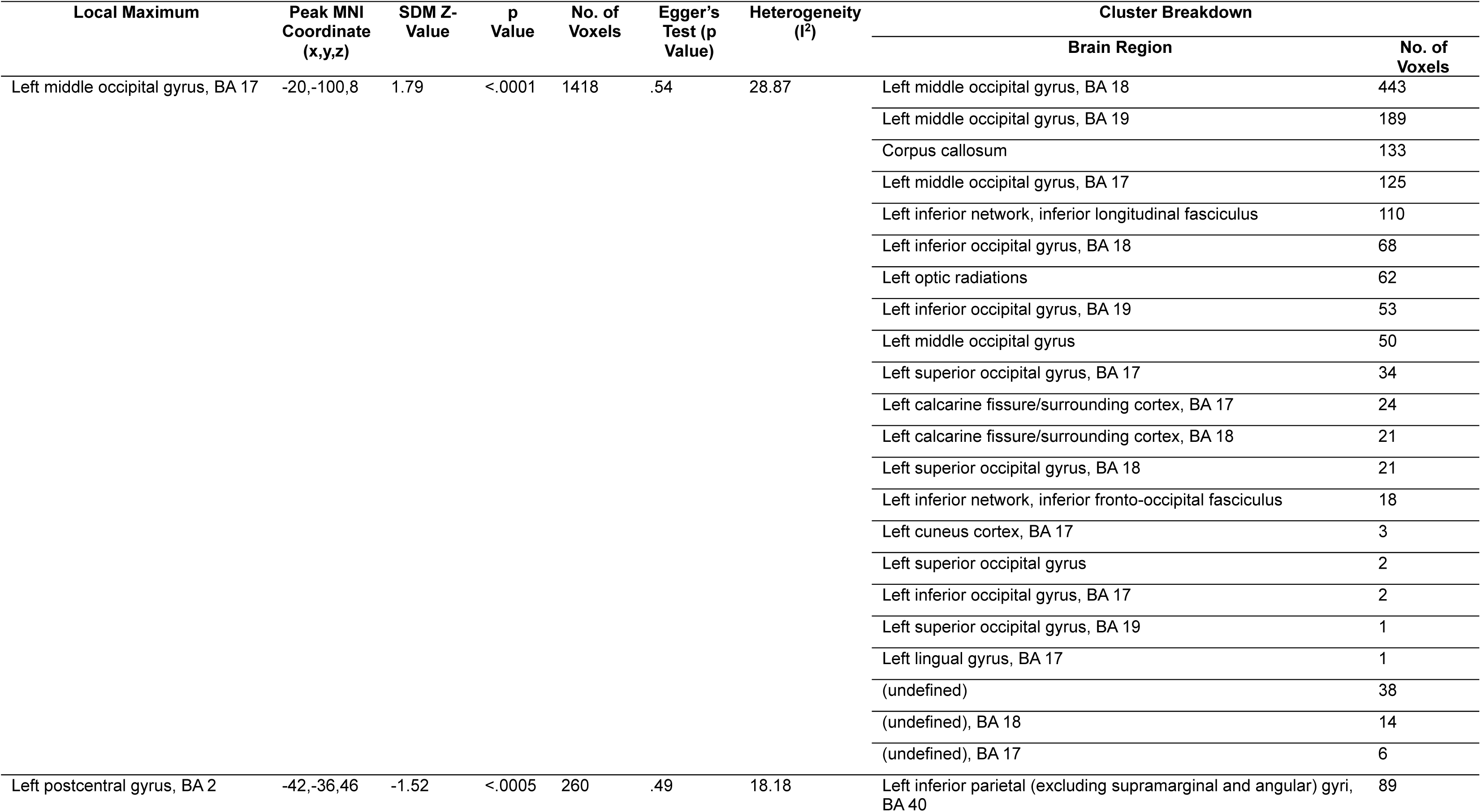
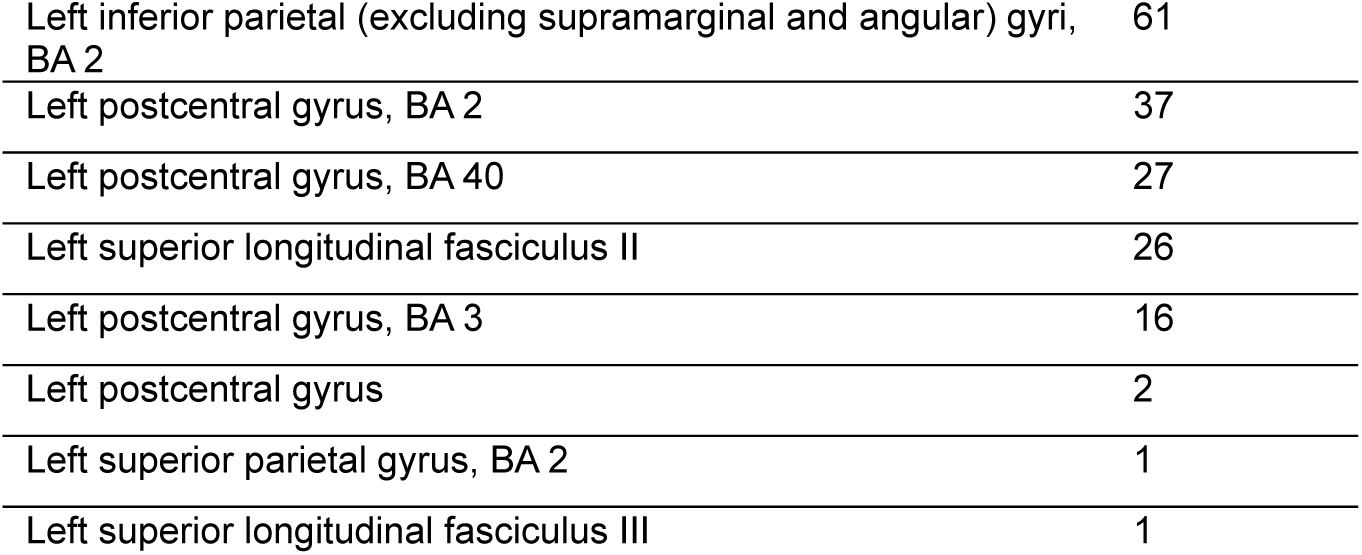
Meta-analysis of fMRI activity differences between individuals with current, chronic AN-r and HCs. Abbreviations: fMRI = functional magnetic imaging; HCs = healthy controls; AN-r = restrictive anorexia nervosa; SDM = seed-based differential mapping; MNI =Montreal Neurological Institute; BA =Brodmann area.

| Local Maximum | Peak MNI Coordinate (x,y,z) | SDM Z-Value | p Value | No. of Voxels | Egger's Test (p Value) | Heterogeneity (I<sup>2</sup>) | Cluster Breakdown |  |
| --- | --- | --- | --- | --- | --- | --- | --- | --- |
|  |  |  |  |  |  |  | Brain Region | No. of Voxels |
| Left middle occipital gyrus, BA 17 | -20,-100,8 | 1.79 | <.0001 | 1418 | .54 | 28.87 | Left middle occipital gyrus, BA 18 | 443 |
|  |  |  |  |  |  |  | Left middle occipital gyrus, BA 19 | 189 |
|  |  |  |  |  |  |  | Corpus callosum | 133 |
|  |  |  |  |  |  |  | Left middle occipital gyrus, BA 17 | 125 |
|  |  |  |  |  |  |  | Left inferior network, inferior longitudinal fasciculus | 110 |
|  |  |  |  |  |  |  | Left inferior occipital gyrus, BA 18 | 68 |
|  |  |  |  |  |  |  | Left optic radiations | 62 |
|  |  |  |  |  |  |  | Left inferior occipital gyrus, BA 19 | 53 |
|  |  |  |  |  |  |  | Left middle occipital gyrus | 50 |
|  |  |  |  |  |  |  | Left superior occipital gyrus, BA 17 | 34 |
|  |  |  |  |  |  |  | Left calcarine fissure/surrounding cortex, BA 17 | 24 |
|  |  |  |  |  |  |  | Left calcarine fissure/surrounding cortex, BA 18 | 21 |
|  |  |  |  |  |  |  | Left superior occipital gyrus, BA 18 | 21 |
|  |  |  |  |  |  |  | Left inferior network, inferior fronto-occipital fasciculus | 18 |
|  |  |  |  |  |  |  | Left cuneus cortex, BA 17 | 3 |
|  |  |  |  |  |  |  | Left superior occipital gyrus | 2 |
|  |  |  |  |  |  |  | Left inferior occipital gyrus, BA 17 | 2 |
|  |  |  |  |  |  |  | Left superior occipital gyrus, BA 19 | 1 |
|  |  |  |  |  |  |  | Left lingual gyrus, BA 17 | 1 |
|  |  |  |  |  |  |  | (undefined) | 38 |
|  |  |  |  |  |  |  | (undefined), BA 18 | 14 |
|  |  |  |  |  |  |  | (undefined), BA 17 | 6 |
| Left postcentral gyrus, BA 2 | -42,-36,46 | -1.52 | <.0005 | 260 | .49 | 18.18 | Left inferior parietal (excluding supramarginal and angular) gyri, BA 40 | 89 |

**Table 4: Meta-analysis of fMRI activity differences between individuals with current, chronic AN-r and HCs.**
|  |  |
| --- | --- |
| Left inferior parietal (excluding supramarginal and angular) gyri, BA 2 | 61 |
| Left postcentral gyrus, BA 2 | 37 |
| Left postcentral gyrus, BA 40 | 27 |
| Left superior longitudinal fasciculus II | 26 |
| Left postcentral gyrus, BA 3 | 16 |
| Left postcentral gyrus | 2 |
| Left superior parietal gyrus, BA 2 | 1 |
| Left superior longitudinal fasciculus III | 1 |

**Figure 5:**
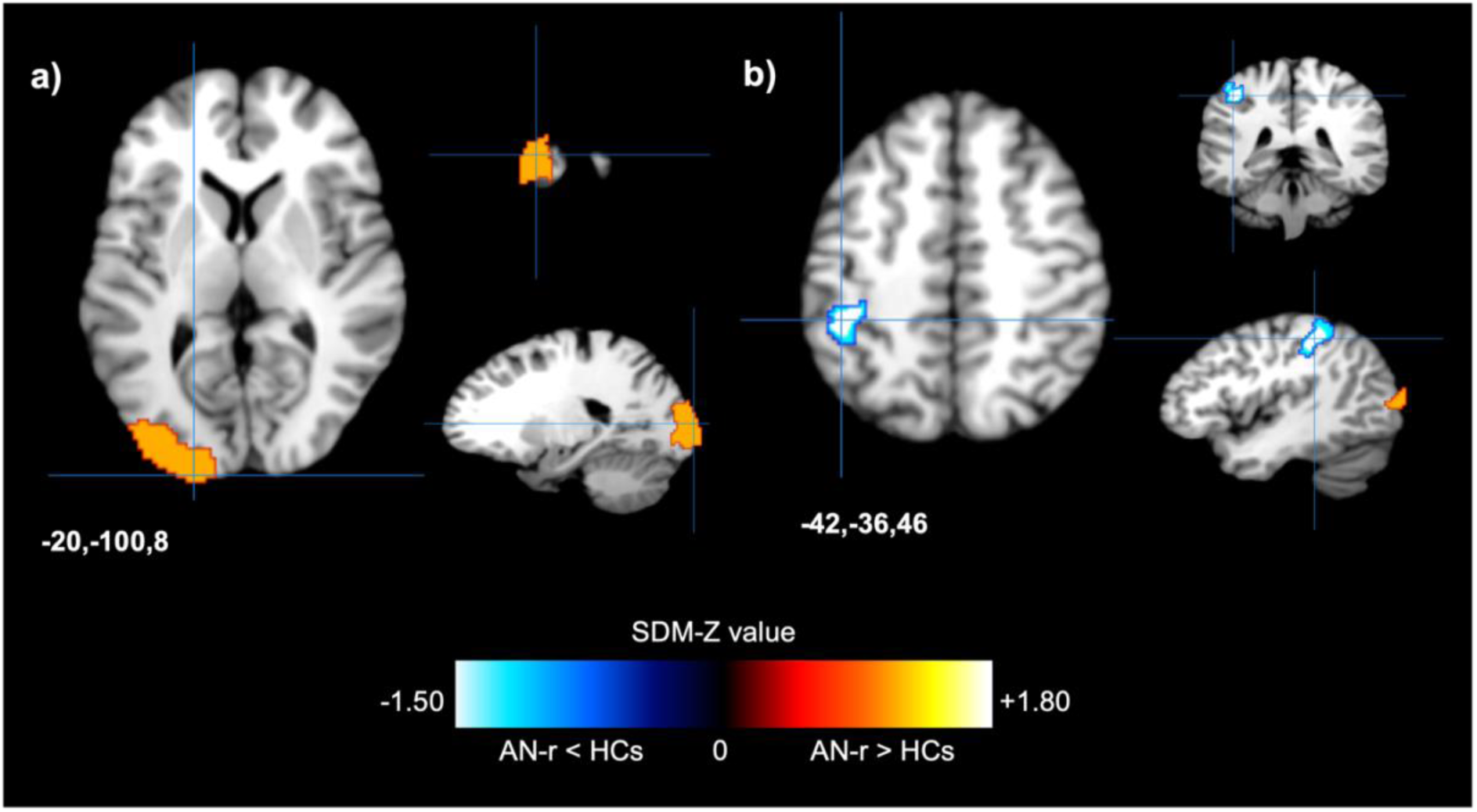
Patients with chronic AN-r showed altered neural activity during reward-based tasks compared to HC. **a.** Patients with chronic AN-r showed greater functional activity in the left middle occipital gyrus (peak MNI coordinates x=-20, y=-100, z=8; max SDM-Z = 1.79) than HCs. **b.** Hypoactivity was observed in the left postcentral gyrus (peak MNI coordinates x=-42, y=-36, z=46; max SDM-Z = −1.52) in patients with chronic AN-r compared to HCs. Abbreviations: HCs = healthy controls; AN-r = restrictive anorexia nervosa patients; SDM = seed-based differential mapping; fMRI = functional magnetic resonance imaging.

#### 3.3.3. Recovered Anorexia Nervosa

Analysis of studies including only patients recovered from AN-r revealed three brain regions where neural activity differed between AN-r and HCs during task performance. Individuals recovered from AN-r exhibited hyperactivity in the right anterior thalamic projections compared to HCs (*g* = 0.25±0.03). Analysis also revealed hypoactivity in the left superior longitudinal fasciculus II (*g* = −0.09±0.01) and left median network (*g* = −0.10±0.02) in recovered individuals compared to HCs (**Table 5**; **Figure 6**).

**Table 5:**
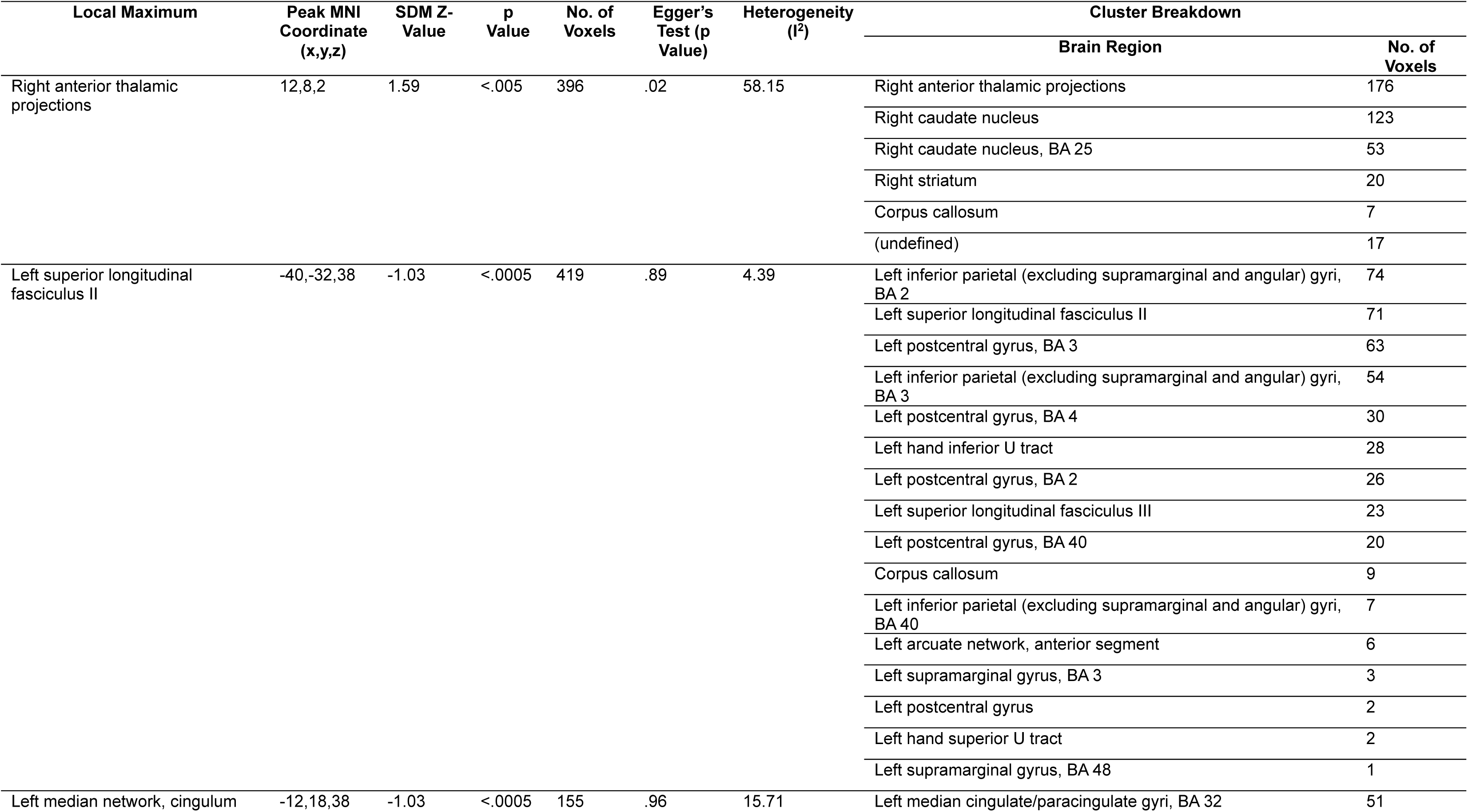
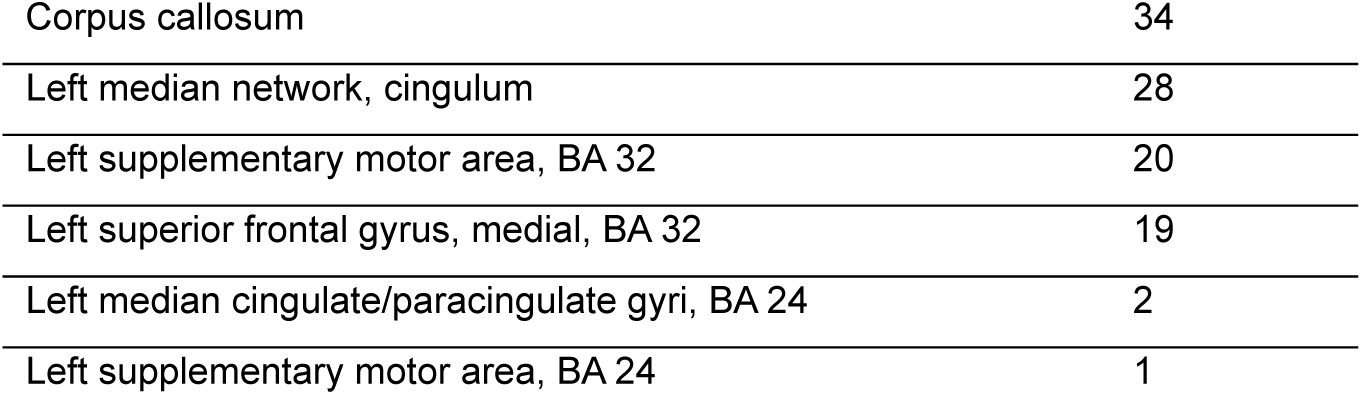
Meta-analysis of fMRI activity differences between individuals recovered from AN-r and HCs. Abbreviations: fMRI = functional magnetic imaging; HCs = healthy controls; AN-r = restrictive anorexia nervosa; SDM = seed-based differential mapping; MNI =Montreal Neurological Institute; BA =Brodmann area.

| Local Maximum | Peak MNI Coordinate (x,y,z) | SDM Z-Value | p Value | No. of Voxels | Egger's Test (p Value) | Heterogeneity (I<sup>2</sup>) | Cluster Breakdown |  |
| --- | --- | --- | --- | --- | --- | --- | --- | --- |
|  |  |  |  |  |  |  | Brain Region | No. of Voxels |
| Right anterior thalamic projections | 12,8,2 | 1.59 | <.005 | 396 | .02 | 58.15 | Right anterior thalamic projections | 176 |
|  |  |  |  |  |  |  | Right caudate nucleus | 123 |
|  |  |  |  |  |  |  | Right caudate nucleus, BA 25 | 53 |
|  |  |  |  |  |  |  | Right striatum | 20 |
|  |  |  |  |  |  |  | Corpus callosum | 7 |
|  |  |  |  |  |  |  | (undefined) | 17 |
| Left superior longitudinal fasciculus II | -40,-32,38 | -1.03 | <.0005 | 419 | .89 | 4.39 | Left inferior parietal (excluding supramarginal and angular) gyri, BA 2 | 74 |
|  |  |  |  |  |  |  | Left superior longitudinal fasciculus II | 71 |
|  |  |  |  |  |  |  | Left postcentral gyrus, BA 3 | 63 |
|  |  |  |  |  |  |  | Left inferior parietal (excluding supramarginal and angular) gyri, BA 3 | 54 |
|  |  |  |  |  |  |  | Left postcentral gyrus, BA 4 | 30 |
|  |  |  |  |  |  |  | Left hand inferior U tract | 28 |
|  |  |  |  |  |  |  | Left postcentral gyrus, BA 2 | 26 |
|  |  |  |  |  |  |  | Left superior longitudinal fasciculus III | 23 |
|  |  |  |  |  |  |  | Left postcentral gyrus, BA 40 | 20 |
|  |  |  |  |  |  |  | Corpus callosum | 9 |
|  |  |  |  |  |  |  | Left inferior parietal (excluding supramarginal and angular) gyri, BA 40 | 7 |
|  |  |  |  |  |  |  | Left arcuate network, anterior segment | 6 |
|  |  |  |  |  |  |  | Left supramarginal gyrus, BA 3 | 3 |
|  |  |  |  |  |  |  | Left postcentral gyrus | 2 |
|  |  |  |  |  |  |  | Left hand superior U tract | 2 |
|  |  |  |  |  |  |  | Left supramarginal gyrus, BA 48 | 1 |
| Left median network, cingulum | -12,18,38 | -1.03 | <.0005 | 155 | .96 | 15.71 | Left median cingulate/paracingulate gyri, BA 32 | 51 |

**Table 5: Meta-analysis of fMRI activity differences between individuals recovered from AN-r and HCs.**
|  |  |
| --- | --- |
| Corpus callosum | 34 |
| Left median network, cingulum | 28 |
| Left supplementary motor area, BA 32 | 20 |
| Left superior frontal gyrus, medial, BA 32 | 19 |
| Left median cingulate/paracingulate gyri, BA 24 | 2 |
| Left supplementary motor area, BA 24 | 1 |

**Figure 6:**
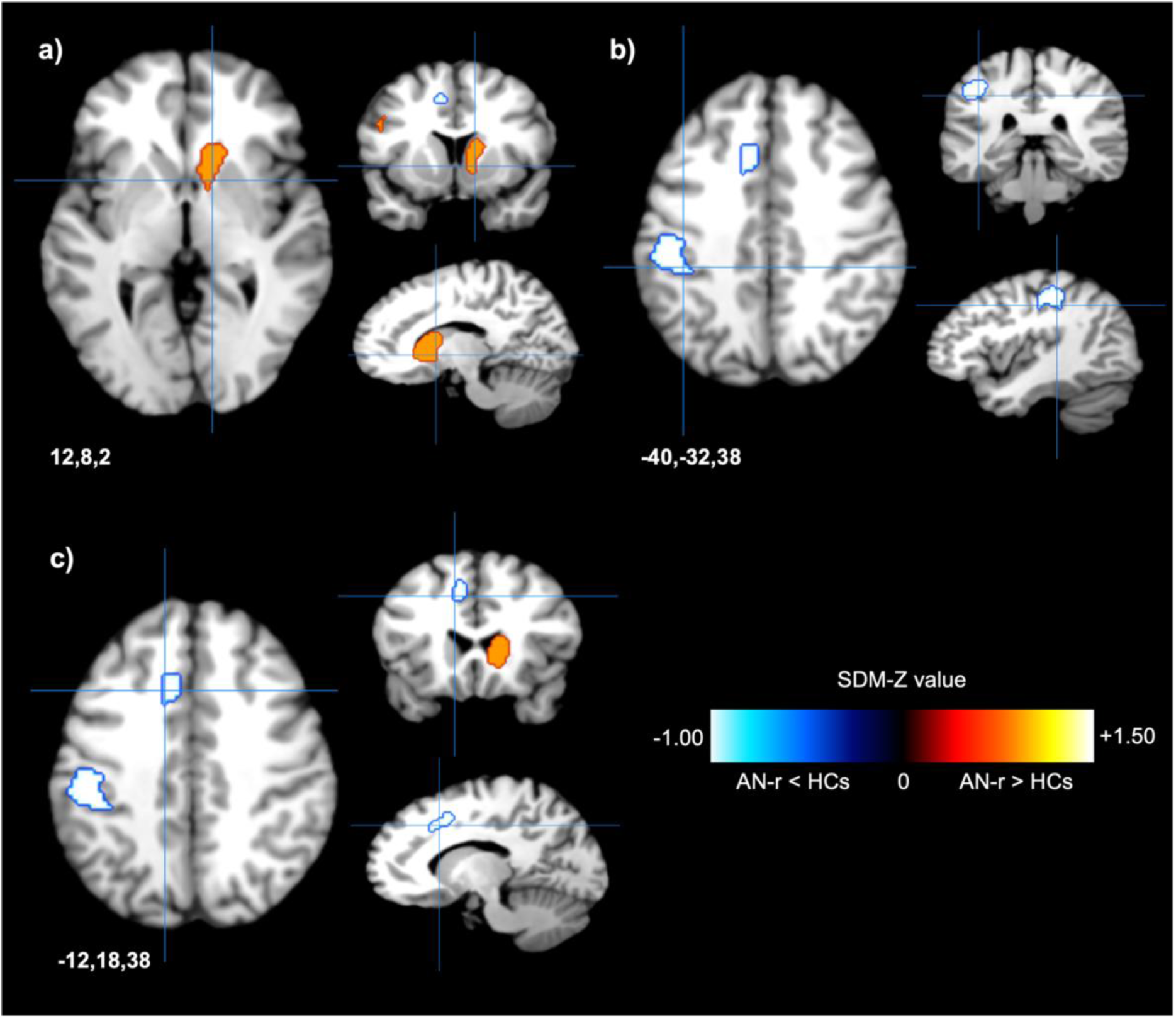
Patients recovered from AN-r showed altered activity during reward-based tasks compared to HC. **a.** Individuals recovered from AN-r exhibited hyperactivity in the right anterior thalamic projections (peak MNI coordinates x=12, y=8, z=2; max SDM-Z = 1.59) **b.** Patients recovered from AN-r showed decreased functional activity in the left superior longitudinal fasciculus II (peak MNI coordinates x=-40, y=-32, z=38; max SDM-Z = - 1.03) compared to HCs. **c.** Hypoactivity was observed in the left median network (cingulum) (peak MNI coordinates x=-12, y=18, z=38; max SDM-Z = −1.03) in patients recovered from AN-r compared to HCs. Abbreviations: HCs = healthy controls; AN-r = restrictive anorexia nervosa patients; SDM = seed-based differential mapping; fMRI = functional magnetic resonance imaging.

### 3.4. Heterogeneity and Publication Bias

In the task-based meta-analyses, of the 18 brain regions with altered reward-related activity identified, only one showed publication bias (Egger’s p < .05: right anterior thalamic projections, recovered AN-r, p = .02) and only two exhibited significant heterogeneity (I^2^ > 50%: left insula BA 48, acute AN-r, 63.04%; right anterior thalamic projections, recovered AN-r, 58.15%) between the included studies.

### 3.5. Meta-Regression Analyses

Meta-regression analysis exploring the associations between the analytic results and age revealed that age was not associated with AN-related brain activity changes (p = .29). Similarly, illness duration did not significantly influence the effect sizes of the studies (p = .37). Effect size was also not influenced by AAO (p = .17) or year of publication (p = .63) (**Figure 7**). Based on these data, it appears that clinical characteristics are not predictors of differences in true effect sizes.

**Figure 7:**
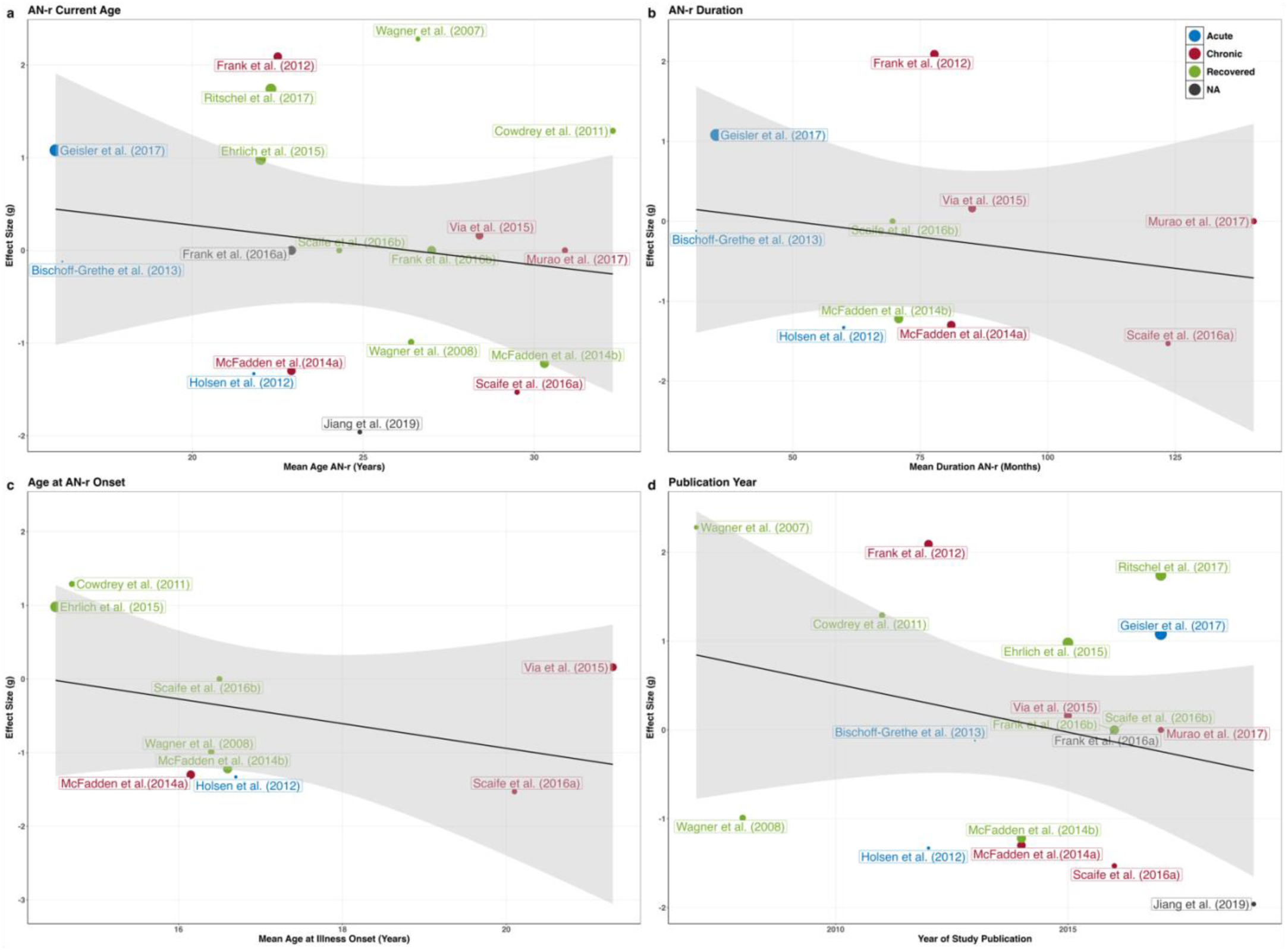
Results of meta-regression analyses. Figures a-d show the relationship between effect size (g) and the clinical characteristics of interest (95% CIs). Point size is proportional to study sample size. **a.** patient age did not influence the effect sizes of the included studies (p = .29). **b.** Illness duration did not influence the effect sizes of the included studies (p = .37). **c.** Age at illness onset was not related to effect size (p = .36). **d.** Publication year was not correlated with effect size (p = 0.63).

### 3.6. Exploratory Analyses

Exploratory analyses were conducted (i) excluding studies reporting comorbidities, (ii) excluding studies reporting the use of psychotropic medications, (ii) taking task stimulus as a covariate.

#### 3.6.1. Excluding Studies Reporting Psychiatric Comorbidities (n = 8)

When excluding studies reporting comorbid conditions, BMI differences between AN-r (*x̃* = 18.8) and HCs (*x̃* = 21.5) remained significant (U = 1, p = .002). Mean age did not differ between individuals with AN-r and HCs (t_(11.5)_ = - 0.05, p = .96). Analysis of these studies revealed hyperactivity in the right precentral gyrus (*g* = 0.18±0.02), left middle frontal gyrus (*g* = 0.18±0.02), left supplementary motor area (*g* = 0.17±0.02) and right anterior thalamic projections (*g* = 0.28±0.05) in patients compared to HCs during reward-related tasks. In contrast, patients with AN-r exhibited hypoactivity in the right precuneus (*g* = −0.12±0.02) compared to HCs (**Table 6**, **Figure 8**). There was no difference in effect size between this exploratory analysis and that including all identified studies (t_1_= 0.51, p = .70). This indicates that findings are robust and alterations in reward-related neural activity are specifically associated with AN-r, not psychiatric comorbidities.

**Table 6:**
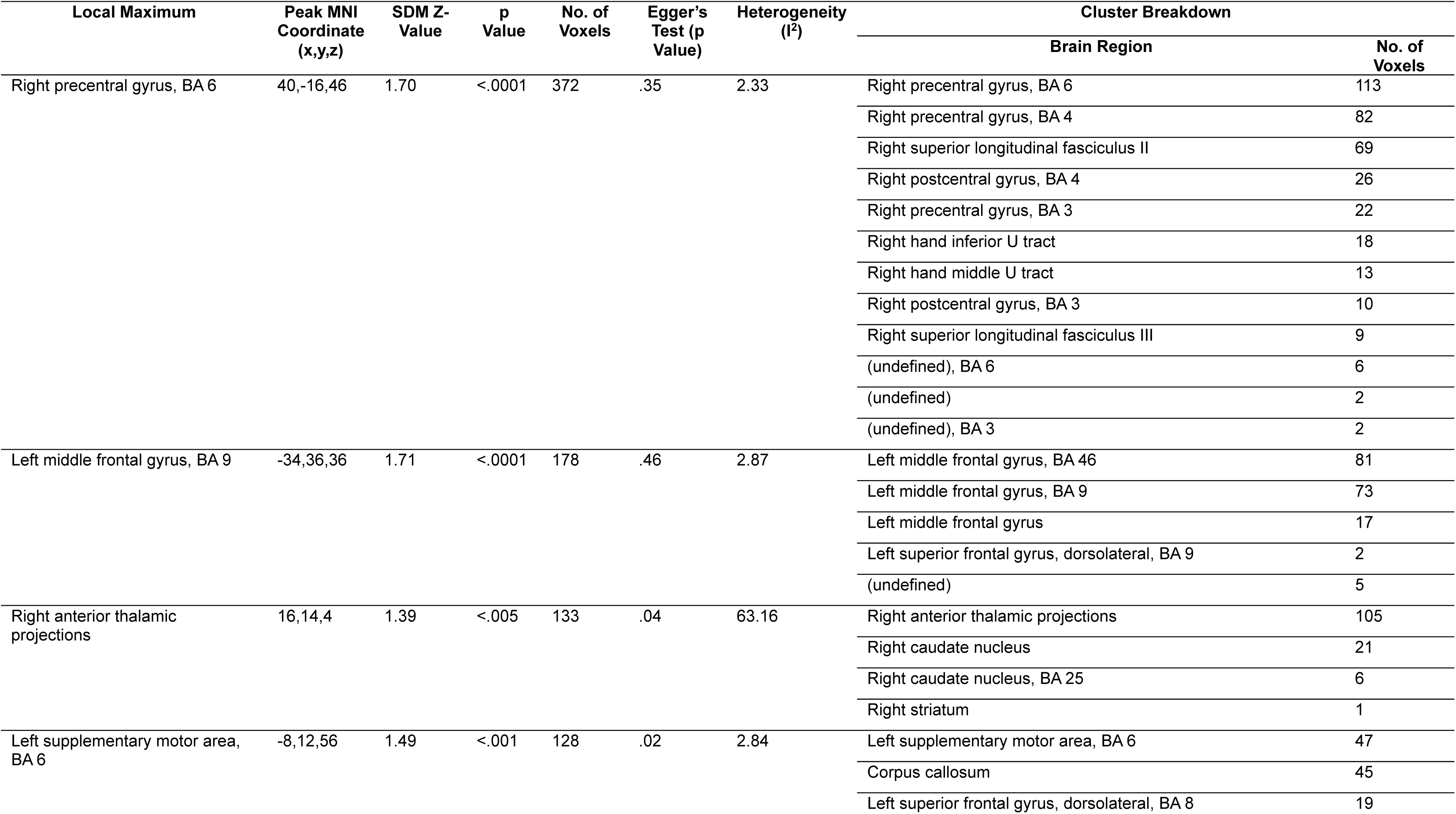
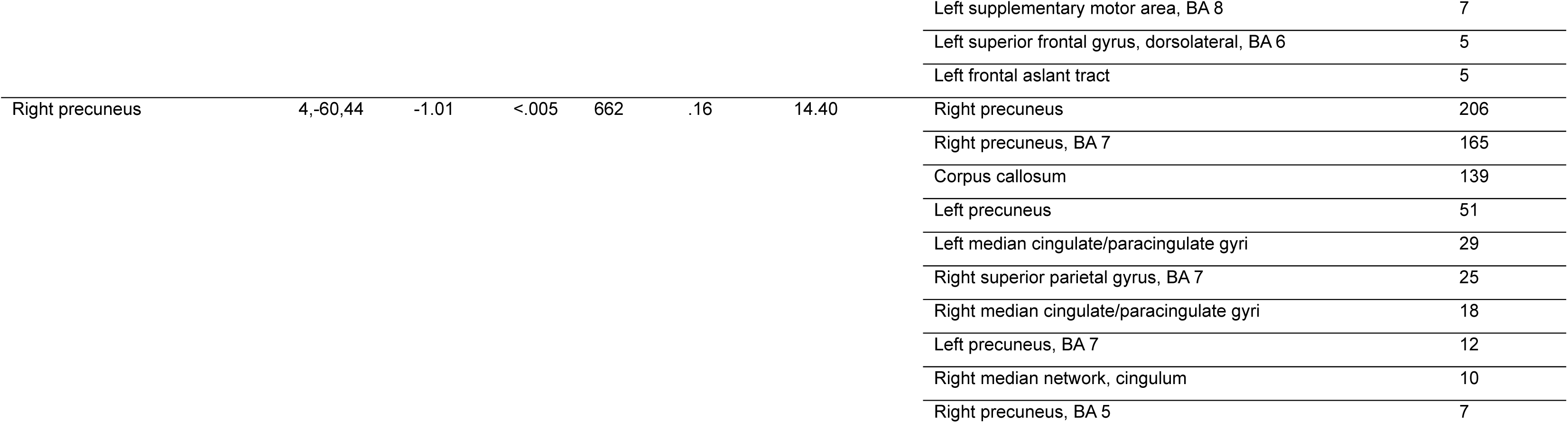
Meta-analysis of fMRI activity differences between individuals with AN-r and HCs, excluding studies reporting psychiatric comorbidities. Abbreviations: fMRI = functional magnetic imaging; HCs = healthy controls; AN-r = restrictive anorexia nervosa; SDM = seed-based differential mapping; MNI =Montreal Neurological Institute; BA =Brodmann area.

**Figure 8:**
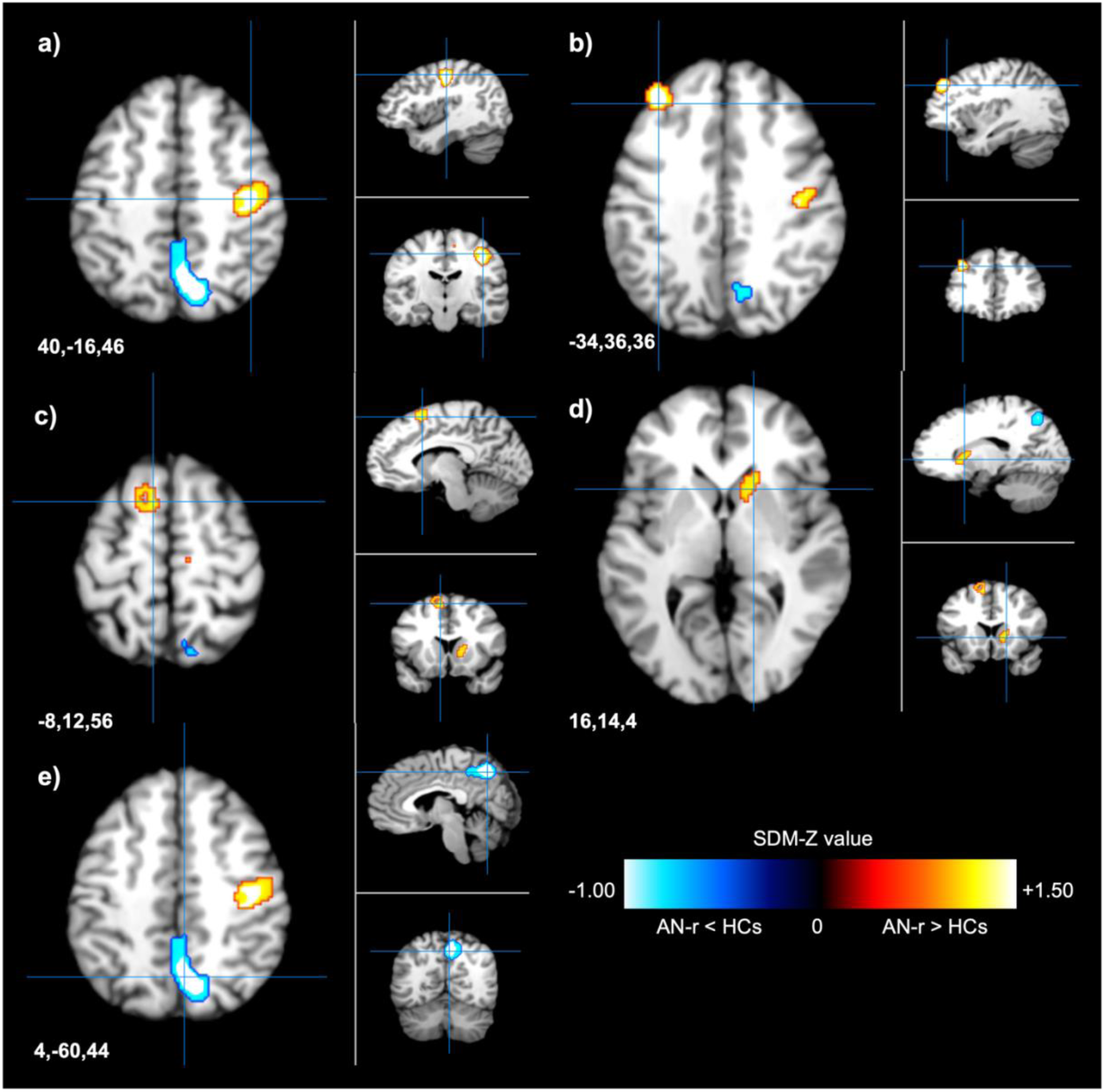
AN-r patients without psychiatric comorbidities exhibited hyperactivity during reward-based tasks compared to HCs. **a.** Patients with AN-r showed greater functional activity in the right precentral gyrus (peak MNI coordinates x=40, y=-16, z=46; max SDM-Z = 1.70) than HCs. **b.** Hyperactivity was observed in the left middle frontal gyrus (peak MNI coordinates x=-34, y=36, z=36, max SDM-Z = 1.71) in patients without psychiatric comorbidities compared to controls **c.** Patients with AN-r showed greater functional activity in the left supplementary motor area (peak MNI coordinates x=-8, y=12, z=56; max SDM-Z = 1.49) than HCs. **d.** Patients with AN-r exhibited hyperactivity in the right anterior thalamic projections (peak MNI coordinates x=16, y=14, z=4; max SDM-Z = 1.39) compared to HCs. **e.** Patients with AN-r exhibited lower functional activity in the right precuneus (peak MNI coordinates x=4, y=-60, z=44; max SDM-Z = −1.01) than HCs. Abbreviations: HCs = healthy controls; AN-r = restrictive anorexia nervosa patients; SDM = seed-based differential mapping; fMRI = functional magnetic resonance imaging.

#### 3.6.2. Excluding Medicated Patients (n = 8)

When patients taking psychoactive medications where excluded, there remained a difference in BMI between patients and controls (t_(8)_ = - 2.53, p = .04). Mean age did not differ between patients and HCs (t_(13)_ = 0.42, p = .68). Analysis using SDM-PSI revealed hyperactivity in the left supplementary motor area (*g* = 0.25±0.02) and right anterior thalamic projections (*g* = 0.33±0.03) in patients compared to HCs during reward-related tasks. In comparison, AN-r patients exhibited hypoactivation in the right hippocampus (*g* = - 0.13±0.03) (**Table 7**, **Figure 9**). Exclusion of medicated patients and their relative controls reliably produced the same results as obtained for the entire sample (t_1_= 0.79, p = .57). This not only indicates that our main findings are robust, but it also suggests that whilst currently available psychiatric medications may aid weight restoration in AN-r (Frank, 2020), they are ineffective in ‘normalising’ reward-related activity in AN-r patients.

**Table 7:**
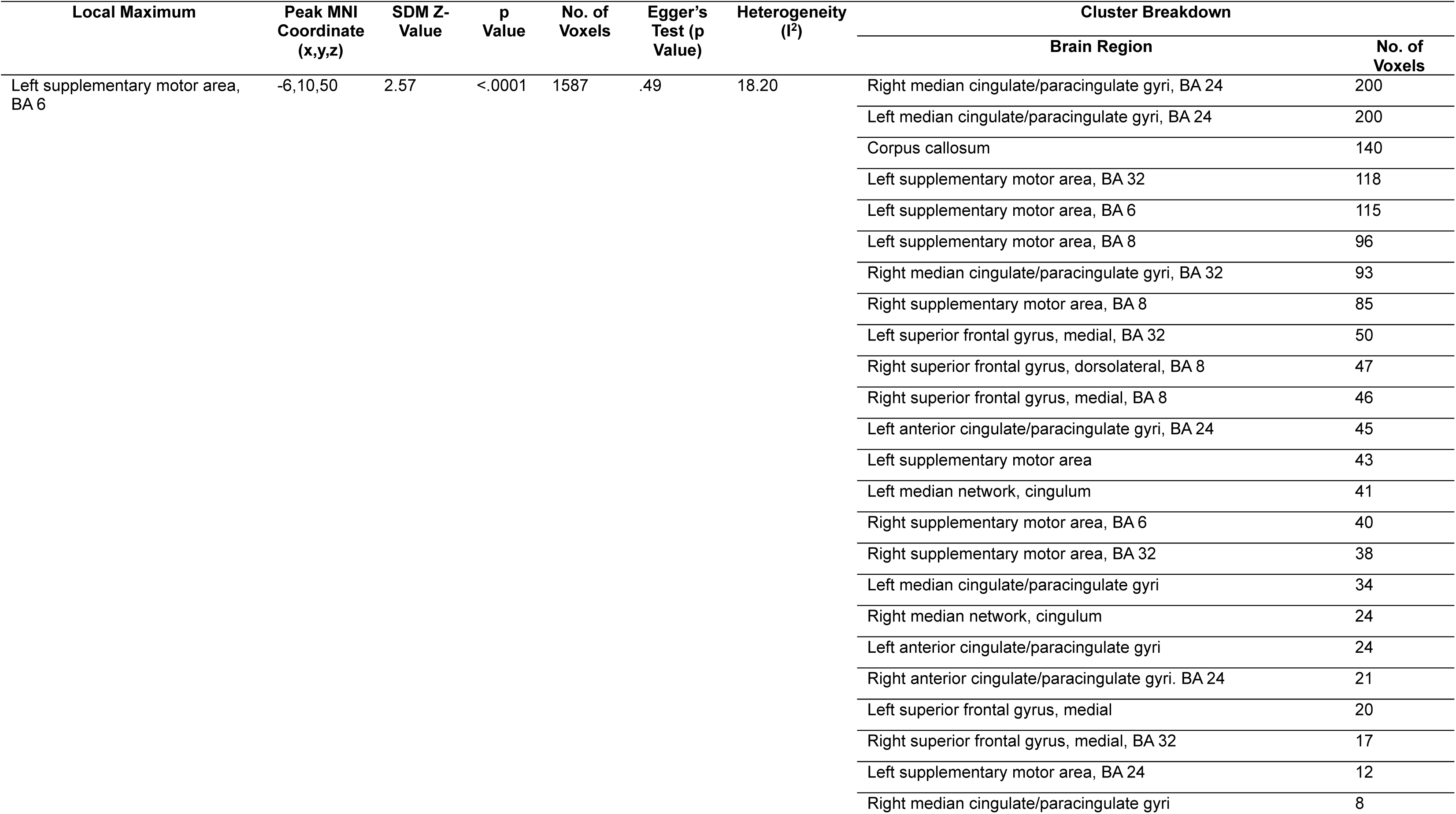
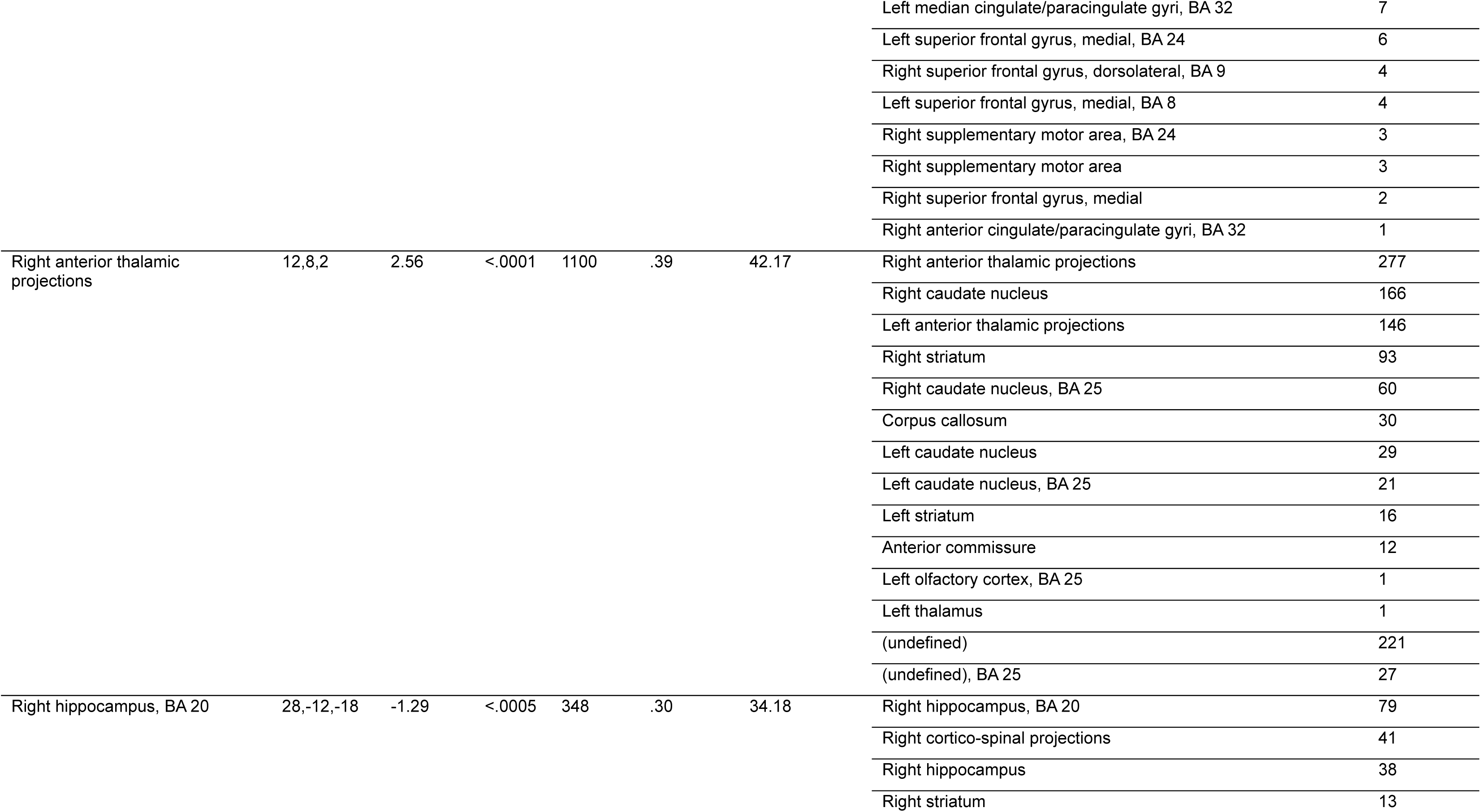
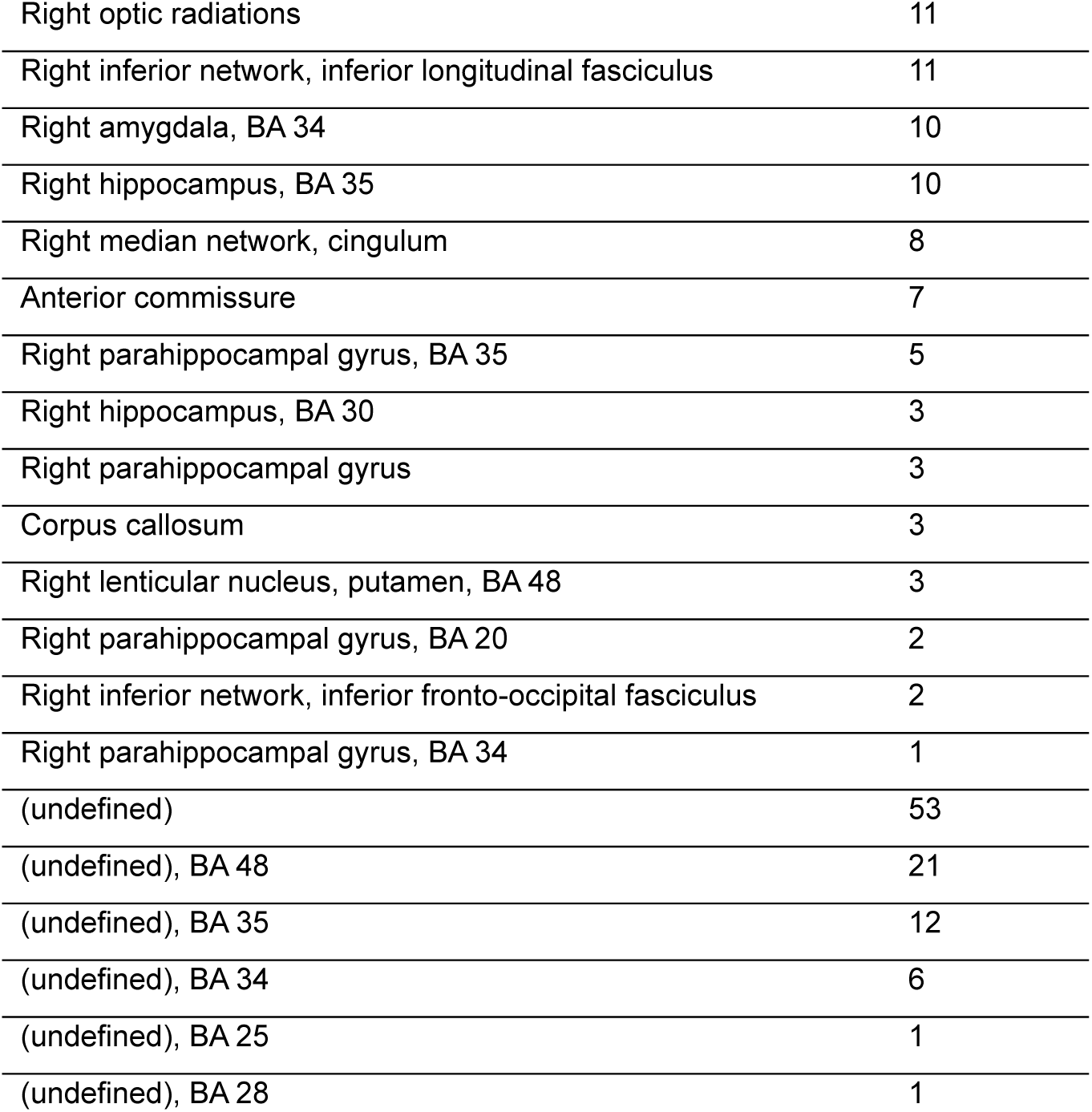
Meta-analysis of fMRI activity differences between individuals with AN-r and HCs, excluding studies reporting the use of psychotropic medication. Abbreviations: fMRI = functional magnetic imaging; HCs = healthy controls; AN-r = restrictive anorexia nervosa; SDM = seed-based differential mapping; MNI =Montreal Neurological Institute; BA =Brodmann area.

**Figure 9:**
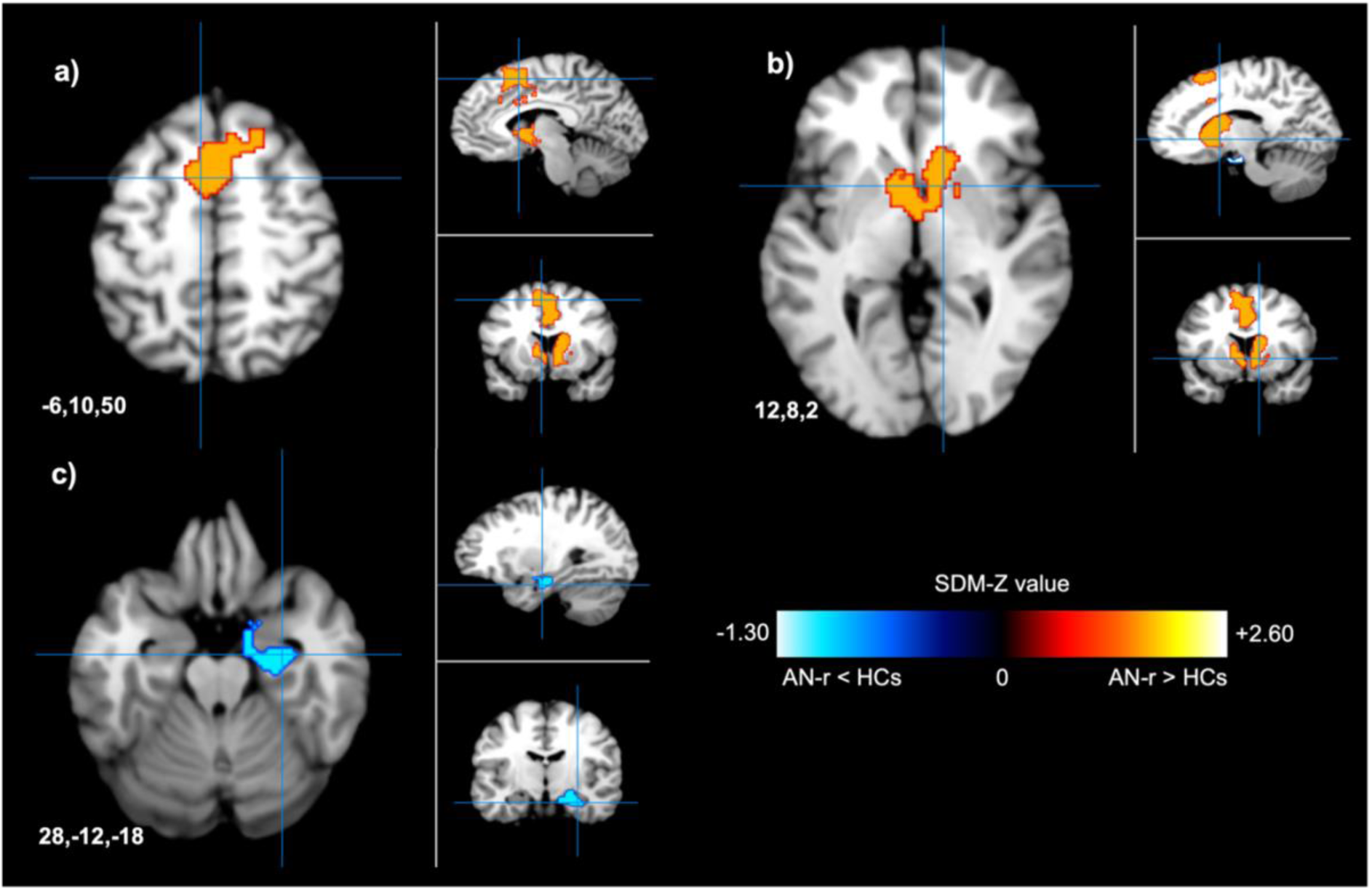
AN-r patients not taking any psychiatric medication exhibited altered activity during reward-based tasks compared to HCs. **a.** Patients with AN-r showed greater functional activity in the left supplementary motor area (peak MNI coordinates x=-6, y=10, z=50; max SDM-Z = 2.57) than HCs. **b.** Hyperactivity was observed in the right anterior thalamic projections (peak MNI coordinates x=12, y=8, z=2, max SDM-Z = 2.56) in patients not taking psychiatric medication compared to controls **c.** Patients with AN-r exhibited lower functional activity in the right hippocampus (peak MNI coordinates x=28, y=-12, z=-18; max SDM-Z = −1.29) than HCs. Abbreviations: HCs = healthy controls; AN-r = restrictive anorexia nervosa patients; SDM = seed-based differential mapping; fMRI = functional magnetic resonance imaging.

#### 3.6.3. Taking Task as a Covariate

When considering task as a covariate in the model, no differences were observed in the BMI of either patients (F_(2,15)_ = 0.18, p = .83) or controls (F_(2,15)_ = 0.71, p = .51) between task groups. The same was true for age, with neither patient age (F_(2,15)_ = 0.60, p = .56) or control age (F_(2,15)_ = 1.29, p = .30) differing between tasks. Meta-analysis with task as a covariate revealed several brain regions, including the striatum, precuneus and insula, in which reward-related activity differed (**Table 8**, **Figure 10**). There was also a significant difference in effect size between task subgroups (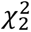 = 7.02, df = 2, p = .03) (**Figure 3**). This indicates that the direction and magnitude of effect is strongly influenced by the choice of task.

**Table 8:** Meta-analysis of fMRI activity differences between individuals with AN-r and HCs, taking task stimulus as a covariate in the model. Abbreviations: fMRI = functional magnetic imaging; HCs = healthy controls; AN-r = restrictive anorexia nervosa; SDM = seed-based differential mapping; MNI =Montreal Neurological Institute; BA =Brodmann area. Note: Direction of effect is not given since this is dependent on which task stimulus is taken as the baseline.

| Local Maximum | Peak MNI Coordinate (x,y,z) | SDM Z-Value | p Value | No. of Voxels | Egger's Test (p Value) | Heterogeneity (I<sup>2</sup>) | Cluster Breakdown |  |
| --- | --- | --- | --- | --- | --- | --- | --- | --- |
|  |  |  |  |  |  |  | Brain Region | No. of Voxels |
| Right inferior parietal (excluding supramarginal and angular) gyri, BA 39 | 56,-56,44 | 2.19 | <.001 | 240 | .46 | 15.95 | Right inferior parietal (excluding supramarginal and angular) gyri, BA 40 | 113 |
|  |  |  |  |  |  |  | Right angular gyrus, BA 39 | 54 |
|  |  |  |  |  |  |  | Right inferior parietal (excluding supramarginal and angular) gyri, BA 39 | 44 |
|  |  |  |  |  |  |  | Right angular gyrus, BA 40 | 12 |
|  |  |  |  |  |  |  | Right supramarginal gyrus, BA 40 | 11 |
|  |  |  |  |  |  |  | Right inferior parietal (excluding supramarginal and angular) gyri | 4 |
|  |  |  |  |  |  |  | Right angular gyrus, BA 48 | 2 |
| Right inferior occipital gyrus, BA 19 | 38,-80,-10 | 1.88 | <.005 | 140 | .30 | 36.20 | Right inferior occipital gyrus, BA 19 | 98 |
|  |  |  |  |  |  |  | Right inferior network, inferior longitudinal fasciculus | 36 |
|  |  |  |  |  |  |  | Right fusiform gyrus, BA 19 | 6 |
| Left anterior cingulate/paracingulate gyri, BA 11 | -2,30,-10 | 1.84 | <.005 | 136 | .39 | 18.64 | Left superior frontal gyrus, medial orbital, BA 11 | 41 |
|  |  |  |  |  |  |  | Right superior frontal gyrus, medial orbital, BA 11 | 37 |
|  |  |  |  |  |  |  | Left anterior cingulate/paracingulate gyri, BA 11 | 37 |
|  |  |  |  |  |  |  | Right anterior cingulate/paracingulate gyri, BA 11 | 10 |
|  |  |  |  |  |  |  | Left gyrus rectus, BA 11 | 9 |
|  |  |  |  |  |  |  | Right gyrus rectus | 1 |
|  |  |  |  |  |  |  | Left superior frontal gyrus, medial orbital | 1 |
| Left inferior frontal gyrus, opercular part, BA 44 | -44,6,28 | 2.27 | <.0005 | 104 | .31 | 3.49 | Left inferior frontal gyrus, opercular part, BA 44 | 47 |
|  |  |  |  |  |  |  | Left precentral gyrus, BA 44 | 27 |
|  |  |  |  |  |  |  | Left inferior frontal gyrus, triangular part, BA 48 | 10 |
|  |  |  |  |  |  |  | Left inferior frontal gyrus, opercular part, BA 48 | 10 |
|  |  |  |  |  |  |  | Left frontal inferior longitudinal fasciculus | 4 |
|  |  |  |  |  |  |  | Left inferior frontal gyrus, triangular part, BA 44 | 3 |
|  |  |  |  |  |  |  | Left superior longitudinal fasciculus III | 2 |
|  |  |  |  |  |  |  | Left inferior frontal gyrus, opercular part | 1 |
| Right middle frontal gyrus, orbital part, BA 10 | 36,58,-4 | 2.03 | <.005 | 97 | .76 | 7.46 | Right middle frontal gyrus, orbital part, BA 47 | 32 |
|  |  |  |  |  |  |  | Right superior frontal gyrus, orbital part, BA 11 | 12 |
|  |  |  |  |  |  |  | Right middle frontal gyrus, orbital part, BA 10 | 11 |
|  |  |  |  |  |  |  | Right middle frontal gyrus, orbital part, BA 11 | 9 |
|  |  |  |  |  |  |  | Right anterior thalamic projections | 8 |
|  |  |  |  |  |  |  | Right middle frontal gyrus, BA 10 | 6 |
|  |  |  |  |  |  |  | Right middle frontal gyrus, orbital part, BA 46 | 6 |
|  |  |  |  |  |  |  | Right middle frontal gyrus, BA 46 | 5 |
|  |  |  |  |  |  |  | Right striatum | 3 |
|  |  |  |  |  |  |  | Right middle frontal gyrus, orbital part | 2 |
|  |  |  |  |  |  |  | Right middle frontal gyrus, BA 47 | 1 |
|  |  |  |  |  |  |  | Right fronto-marginal tract | 1 |
|  |  |  |  |  |  |  | Right middle frontal gyrus, BA 11 | 1 |
| Left striatum | -20,8,-10 | 1.30 | <.0005 | 672 | .94 | 7.29 | Left striatum | 190 |
|  |  |  |  |  |  |  | Left lenticular nucleus, putamen, BA 48 | 133 |
|  |  |  |  |  |  |  | Left amygdala, BA 34 | 58 |
|  |  |  |  |  |  |  | Anterior commissure | 35 |
|  |  |  |  |  |  |  | Left olfactory cortex, BA 48 | 20 |
|  |  |  |  |  |  |  | Left pons | 16 |
|  |  |  |  |  |  |  | Left lenticular nucleus, putamen, BA 25 | 11 |
|  |  |  |  |  |  |  | Left cortico-spinal projections | 9 |
|  |  |  |  |  |  |  | Left lenticular nucleus, putamen, BA 11 | 6 |
|  |  |  |  |  |  |  | Left anterior thalamic projections | 6 |
|  |  |  |  |  |  |  | Left hippocampus, BA 34 | 5 |
|  |  |  |  |  |  |  | Left inferior network, inferior fronto-occipital fasciculus | 3 |
|  |  |  |  |  |  |  | Left hippocampus | 3 |
|  |  |  |  |  |  |  | Left amygdala | 2 |
|  |  |  |  |  |  |  | Left olfactory cortex, BA 25 | 1 |
|  |  |  |  |  |  |  | Left amygdala, BA 48 | 1 |
|  |  |  |  |  |  |  | Left olfactory cortex | 1 |
|  |  |  |  |  |  |  | Left lenticular nucleus, putamen, BA 34 | 1 |
|  |  |  |  |  |  |  | Left inferior network, uncinate fasciculus | 1 |
|  |  |  |  |  |  |  | Left olfactory cortex, BA 11 | 1 |
|  |  |  |  |  |  |  | Left lenticular nucleus, putamen | 1 |
|  |  |  |  |  |  |  | Left olfactory cortex, BA 34 | 1 |
|  |  |  |  |  |  |  | (undefined), BA 48 | 90 |
|  |  |  |  |  |  |  | (undefined), BA 34 | 44 |
|  |  |  |  |  |  |  | (undefined), BA 25 | 16 |
|  |  |  |  |  |  |  | (undefined) | 15 |
|  |  |  |  |  |  |  | (undefined), BA 11 | 2 |
| Right precuneus | 4,-62,34 | 1.31 | <.0005 | 421 | .94 | 1.38 | Left precuneus | 178 |
|  |  |  |  |  |  |  | Right precuneus | 108 |
|  |  |  |  |  |  |  | Left precuneus, BA 7 | 31 |
|  |  |  |  |  |  |  | Left precuneus, BA 23 | 23 |
|  |  |  |  |  |  |  | Right precuneus, BA 7 | 20 |
|  |  |  |  |  |  |  | Left cuneus cortex | 20 |
|  |  |  |  |  |  |  | Right median network, cingulum | 17 |
|  |  |  |  |  |  |  | Right precuneus, BA 23 | 9 |
|  |  |  |  |  |  |  | Left median network, cingulum | 6 |
|  |  |  |  |  |  |  | Right median cingulate/paracingulate gyri, BA 23 | 2 |
|  |  |  |  |  |  |  | Right posterior cingulate gyrus | 2 |
|  |  |  |  |  |  |  | Left posterior cingulate gyrus | 2 |
|  |  |  |  |  |  |  | Right posterior cingulate gyrus, BA 23 | 2 |
|  |  |  |  |  |  |  | Corpus callosum | 1 |
| Left postcentral gyrus, BA 2 | -42,-32,42 | 1.12 | <.005 | 110 | .82 | 7.09 | Left inferior parietal (excluding supramarginal and angular) gyri, BA 2 | 37 |
|  |  |  |  |  |  |  | Left superior longitudinal fasciculus II | 23 |
|  |  |  |  |  |  |  | Left postcentral gyrus, BA 2 | 15 |
|  |  |  |  |  |  |  | Left postcentral gyrus, BA 40 | 14 |
|  |  |  |  |  |  |  | Left inferior parietal (excluding supramarginal and angular) gyri, BA 40 | 13 |
|  |  |  |  |  |  |  | Left inferior parietal (excluding supramarginal and angular) gyri, BA 3 | 4 |
|  |  |  |  |  |  |  | Left postcentral gyrus, BA 3 | 2 |
|  |  |  |  |  |  |  | Left hand inferior U tract, BA 3 | 1 |
|  |  |  |  |  |  |  | Corpus callosum | 1 |
| Left insula, BA 48 | -42,-2,4 | 1.06 | <.005 | 62 | .50 | 0.00 | Left insula, BA 48 | 55 |
|  |  |  |  |  |  |  | Left rolandic operculum, BA 48 | 5 |
|  |  |  |  |  |  |  | Left rolandic operculum | 1 |
|  |  |  |  |  |  |  | Left insula | 1 |

**Figure 10:**
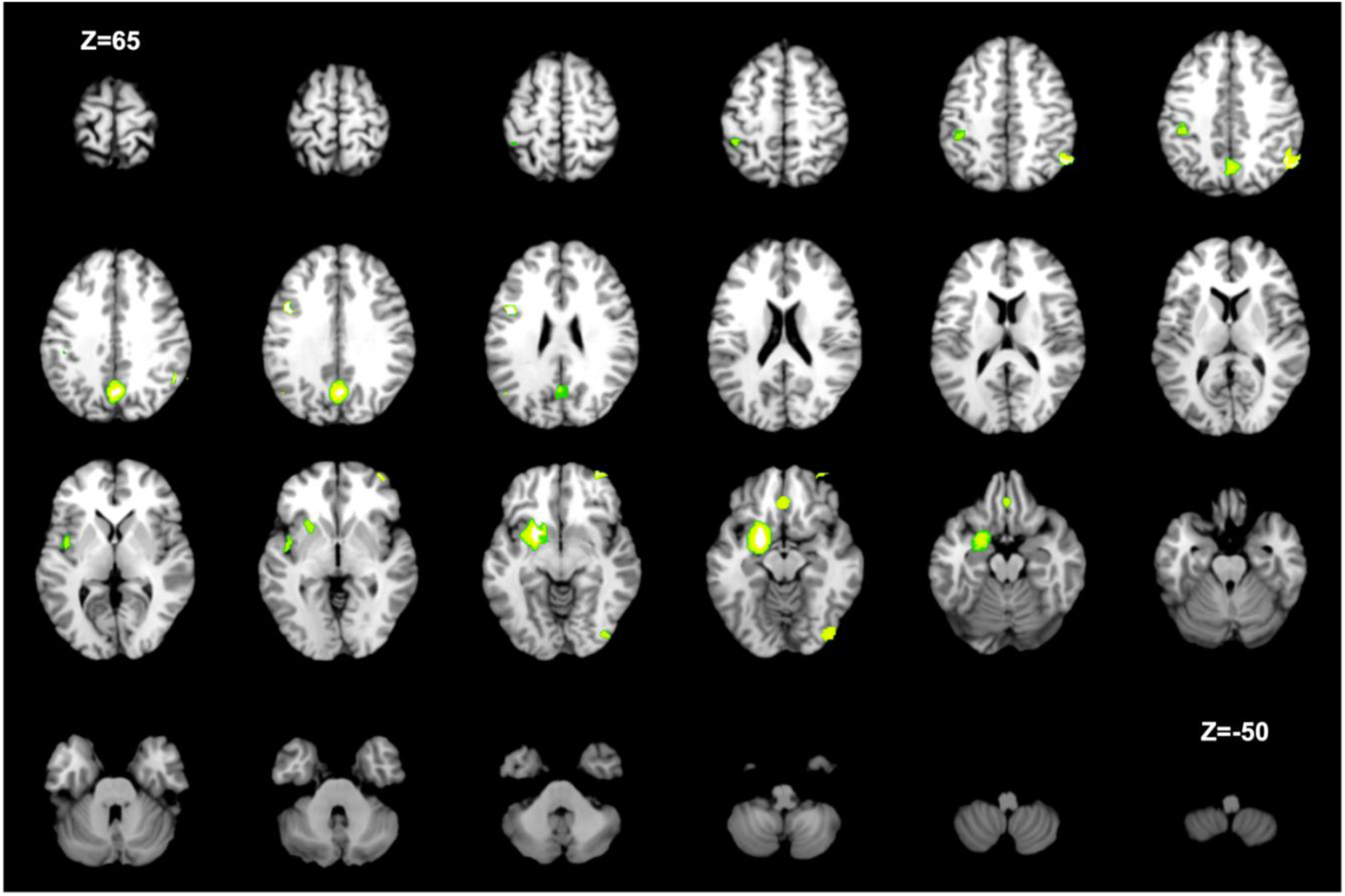
Results of meta-analysis with task stimulus (food, money, body-focused/social) as a variable. Differences in neural activity were observed in the right inferior parietal gyrus, left anterior cingulate/paracingulate gyri, right inferior occipital gyrus, right middle frontal gyrus, left inferior frontal gyrus, left striatum, right precuneus, left postcentral gyrus and left insula as a function of task. Axial images are displayed in MNI Z coordinates. Abbreviations: HCs = healthy controls; AN-r = restrictive anorexia nervosa patients; SDM = seed-based differential mapping; fMRI = functional magnetic resonance imaging. Note: Direction of effect is not given since this is dependent on which task stimulus is taken as the baseline.

## 4. DISCUSSION

In the present study, we sought to determine whether abnormalities in reward-related neural activity are observed in AN-r, and if so, whether they are related to illness stage. We combined the results from 18 neuroimaging datasets and found that AN-r was associated with alterations in reward-related neural activity. This provides support to the widespread notion that altered reward-processing is a biomarker of AN-r (Frank, 2013). However, despite similar effect sizes when subgroup analyses considering illness stage were conducted, it was observed that different brain regions were implicated depending on illness length and recovery status.

### 4.1. AN-r appears to be characterised by reward contamination rather than anhedonia

We observed hyperactivity in the left anterior and median cingulate/paracingulate gyri in acute AN-r compared to HCs. This is consistent with findings from Gaudio et al. (2015) whose study of resting state networks (RSNs) in AN indicated that changes in functional connectivity between the executive control network (ECN) and cingulate cortex may represent an early state-related biomarker of the disease. The cingulate cortex forms part of the ventral limbic circuit which, in conjunction with the ECN, is concerned with inhibitory decision making and reward (Seth & Critchley, 2013). Hyperactivity of the cingulate cortex in acute AN-r therefore suggests difficulty in appropriately regulating behaviours in response to potential reward/punishment. Such findings can be interpreted in line with RCT (Keating, 2010).

As a part of the fronto-striatal network, Keating (2010) proposed that the anterior cingulate cortex (ACC) may represent a key locus for reward-contamination. The ACC is well-documented to be functionally heterogenous and involved in decision making (Pochon et al., 2008), reward (Beck et al., 2006), punishment (Wrase et al., 2007) and reinforcement learning (Kennerley et al., 2006). Furthermore, primate studies have revealed a specific role for the ACC in representing the contingency between actions and outcomes (Hadland et al., 2003). It is therefore plausible that ACC hyperactivity may relate an inability to regulate the conflict between reward and punishment and the maintenance of maladaptive behaviours. Furthermore, our analyses revealed hypoactivity in a region including the right amygdala in acute AN-r. The amygdala has long been recognised to be involved in the integration of sensory information and execution of behaviours dependent on the valence assigned to cues (Everitt et al., 2000; Paton et al., 2006; Smith & Torregrossa, 2021). Altered activity in the amygdala therefore provides further support for a model of reward contamination as opposed to anhedonia in acute AN-r.

Although the precise neurochemistry of altered reward-processing in AN-r is unclear, dopamine (DA) remains the most widely studied reward candidate (Keating et al., 2012). DA release may be modulated by stress, and the hypothalamic-pituitary-adrenal (HPA) axis is involved in stress responding (Lutz et al., 2015). It has been proposed that behaviours linked to AN-r upregulate the HPA axis (Simon et al., 2019), resulting in increased DA release from the ventral striatum (Bergh & Sodersten, 1996). Abnormalities in DA metabolism have been reported in individuals with AN, with one study reporting increases in the concentration of the DA metabolite homovanillic acid (HVA) in the cerebrospinal fluid (CSF) of patients compared to controls (Kaye et al., 1984) Additionally, studies using pavlovian reward tasks in rodents have revealed a role for DA in the motivational component of reward (Smith et al., 2011). Our meta-analysis identified hyperactivity of the right striatum in acute AN-r, suggesting alterations in DA release in the striatum to be a primary biochemical mechanism underlying AN-r symptomatology. The causality of alterations in DA signalling in AN-r is worthy of further study.

Following the framework proposed by Robinson and Berridge (1993) to understand addiction, reward can be dissociated into ‘liking’ and ‘wanting’, where ‘liking’ is considered a hedonic reaction to the pleasure of reward and ‘wanting’ or ‘incentive salience’ associated with the motivation to pursue the ‘rewarding’ stimulus. Berridge et al. (1989) hypothesised that depletion of brain dopamine in rats via a neurochemical lesion would reduce hedonic orofacial expressions elicited by sweet tastes. However, they found that even after dopamine depletion, ‘liking’ reactions were completely normal, despite rats being profoundly aphagic. It is now understood that ‘wanting’ or incentive salience is generated by mesolimbic dopamine originating in the striatum (Frank et al., 2005). In contrast, ‘liking’ is not dependent on dopamine (Berridge & Kringelbach, 2015). Whilst these two neuronal systems usually function in concert, it has been proposed that where a particular behaviour becomes pathological, as in addiction and AN-r, tagging of incentive salience to the behaviour may spur the continued engagement in such behaviours without a concurrent alteration in ‘liking’ responses (Keating et al., 2012). This mechanism may facilitate the maintenance of AN-r, with constant disorder-induced dopamine release reinforcing behaviours that would normally be considered aversive (Cowdrey et al., 2011; Sodersten et al., 2016). In support of this hypothesis, Drewnowski et al. (1987) found that sensory estimates of sweetness and fat content (i.e., ‘liking’) did not differ between AN patients and HCs, but ‘wanting’ for sugar relative to fat did, with AN-r patients showing elevated optimal sugar:fat ratios. It therefore appears plausible that our finding of ACC hyperactivity in acute AN-r and the resulting failure to regulate the conflict between reward and punishment may be a consequence of elevated dopaminergic tone in the striatum (Frank et al., 2005). Dopaminergic signalling however has not been investigated in AN-r patients or HCs during reward processing tasks, and further investigation of RCT as a framework for understanding the key symptoms of AN-r is required.

### 4.2. Interoceptive dysregulation may represent an early marker of general psychopathology

Functional activity of the insula during reward processing further distinguished individuals with acute AN-r from controls in our analysis. This indicates that interoceptive dysregulation may represent an early marker of the disease which contributes to reward contamination (Keating, 2010) and altered subjective responses to stimuli such as food, hunger and physical sensations (Strigo et al., 2013). Through connections with the frontal cortex and hypothalamus, the insula is critically involved in the regulation of appetite and eating (Koh et al., 2003), whilst bidirectional projections to structures of the limbic system are involved in emotion regulation and the integration of thoughts and feelings with sensory experiences (Shelley & Trimble, 2004). In line with the neurobiological hypothesis of AN proposed by Nunn and colleagues (2008), we hypothesise that insula dysfunction is likely a predisposing risk factor, which, when coupled with precipitating factors such as socio-cultural pressures, family complications and puberty, may result in overt psychopathology. Due to its vast connections with frontal, temporal and parietal structures, dysfunction of the insula would be predicted to lead to dysfunction in several brain networks, resulting in the inability to reliably integrate affective and sensory information and failure to inhibit weight and shape concerns (Nunn et al., 2008). Over time, this would lead to the reinforcement of pathological behaviours (Sodersten et al., 2016), reward contamination (Keating, 2010) and disorder progression.

Consistent with this interpretation, individuals with acute AN-r also showed altered activity in the right anterior thalamic projections and right striatum (particularly the dorsal striatum), which has strong connections with the insula (Nunn et al., 2008), and which may serve as a neural marker of general psychopathology (Luo et al., 2019). Traditionally, the dorsal striatum, which includes the caudate nucleus and putamen, was conceptualised as mainly involved in motor control (e.g. DeLong, 1973; Whishaw et al., 1986). However volumetric and functional differences have since been implicated in a variety of neuropsychiatric and neurodevelopmental conditions including schizophrenia (Mitelman et al., 2009), obsessive-compulsive disorder (OCD) (He et al., 2024), attention deficit hyperactivity disorder (ADHD) (Cao et al., 2009) and autism spectrum disorder (ASD) (Lefebvre et al., 2023). The putamen is typically associated with habitual control of behaviour, is overactive in OCD (Gillian, 2021), and appears to exhibit disrupted development in autism (Evans et al., 2024). The cognitive profile of AN-r shares many similarities with that of ASD (Oldershaw et al., 2011) and OCD (Sherman et al., 2006), including cognitive rigidity and repetitive body-focused behaviours. Importantly, in many cases, greater alterations in the putamen have been associated with greater symptom burden and poorer outcomes (Luo et al., 2019). It is therefore possible that altered functional activity in the dorsal striatum in AN-r may be a neural marker of poorer prognosis or habitual control more generally, rather than a specific neural correlate of AN-r. Nonetheless, it is important to note that altered activity in the striatum was not observed in the chronic group, suggesting that these alterations reflect state, rather than trait, markers.

### 4.3. Altered activity in the occipital gyrus may be specifically related to poorer prognosis

Analysis of the chronic group revealed significantly higher functional activity in the left middle occipital gyrus in patients than HCs, with these alterations again tentatively related to long-term outcomes (Boehm et al., 2016). The occipital cortex is a key perceptual area for the early analysis of visual stimuli (Su et al., 2021), leading to the suggestion that individuals with AN-r may experience reward differently at the perceptual level (Boehm et al., 2016; Suchan et al., 2013); namely through body image distortion (BID).

BID is a core diagnostic feature of AN (American Psychiatric Association, 2013), and patients typically experience underweight stimuli as rewarding and overweight stimuli as aversive (Fladung et al., 2013). Body image is a multidimensional construct, consisting of both an attitudinal component arising from a subjective body and shape dissatisfaction (Cornelissen et al., 2013) and a perceptual aspect described as an inability to accurately evaluate body size (Banfield & McCabe, 2002). Whilst the role of attitudinal BID in the aetiology of AN is widely accepted (Keel et al., 2005), the nature of *perceptual* BID remains debated (Cornelissen et al., 2013). Nonetheless, there is some evidence for a positive association between perceptual BID and long-term outcomes (Boehm et al., 2016). For example, in their study of 76 adolescent female AN patients, Boehm et al. (2016) reported that perceptual BID strongly predicted long-term physical, psychological, and psychosocial adjustment as measured using the Morgan-Russell outcome assessment schedule (Morgan & Russell, 1975). Our findings provide further support to this purported association. Hyperactivity in the left middle occipital gyrus was only observed in chronic AN-r, indicating that alterations in the perceptual experience of reward may be specifically related to poorer prognosis and increased illness duration.

An additional consideration is that individuals with AN commonly experience difficulties in recognising the emotions and intentions of others and show an attentional bias toward negative facial expressions (Treasure & Schmidt, 2013). Such impairments often result in social anhedonia (Harrison et al., 2014) and maintenance of the disorder (Tauro et al., 2022). Altered functional activity in the occipital gyrus during reward-based tasks could therefore be associated with deficits in social proficiency (Sweitzer et al., 2018). In their cognitive-interpersonal maintenance model for AN, Schmidt and Treasure (2006) propose that AN is maintained by interpersonal factors including perceived reactions from peers which are complicated by the egosyntonic nature of the disease. This leads to negative emotions, social withdrawal, and ultimately impaired social cognition and social reward processing (Treasure & Schmidt, 2013). Our results provide some support for this model, since of the six studies included in the chronic subgroup meta-analysis, only two reported hyperactivation of visual areas (Frank et al., 2012; Via et al., 2015), and of these, one was the only study in the chronic subgroup to use a social judgement task (Via et al., 2015). This indicates that altered functional activity in the occipital gyrus in chronic AN-r may be related to deficits in social reward processing.

Nonetheless, we must also consider the possibility that our findings of differential reward-based neural activation in acute versus chronic AN-r may be an artefact of the different tasks employed by the original researchers and their associated cognitive demands. In comparison to those included in the chronic analysis which mostly commonly used food-based tasks (67%), most studies of acute AN-r (67%) employed monetary reward tasks. In conjunction with this, our exploratory analyses indicated that the direction and magnitude of effect is strongly influenced by task stimulus. Differences in neural activation may therefore represent differences related to social versus non-social reward rather than illness stage explicitly.

### 4.4. Psychological recovery shows a more protracted course than physical recovery

In recovered individuals, hypoactivity was observed in the left superior longitudinal fasciculus II (SLF) and left median network compared to HCs. The SLF interconnects the frontal lobe with the parietal, occipital and temporal lobes (Naidich et al., 2013) and has been implicated in several cognitive functions including visual working memory (Kinoshita et al., 2016) and language (Madhavan et al., 2014). We therefore suggest that hypoactivity in this region in recovered AN-r likely results from illness-related impairments in cognitive functioning (and thus greater task difficulty) rather than altered reward processing. This interpretation is further supported by the key role of the median network (cingulum bundle) in high-level processing and executive control (Bubb et al., 2018). However, studies which specifically probe task difficulty in individuals recovered from AN-r and HCs will be required to confirm this hypothesis.

Residual hyperactivity in the right anterior thalamic projections and right caudate nucleus in recovered AN-r suggests that despite weight restoration, the underlying psychopathology of AN-r may take longer to ‘normalise’ (Luo et al., 2019). However, it remains unclear whether such alterations in reward-based functional activity are a vulnerability factor which predates illness onset or a result of disease processes.

Considering reports that the grey and white matter reductions observed in AN-r are reversible (at least in non-chronic patients) (Wagner et al., 2006) and not all brain regions are equally affected by volume reductions resulting from acute malnutrition (Scharner & Stengel, 2019), we suggest that reward-based abnormalities should be considered a dynamic state marker of AN-r. It has been well-documented that the frontal-parietal-cingulate network exhibits atrophy that is disproportional with the rest of the brain in AN-r (Bär et al., 2015; McCormick et al., 2008). Since this network is the main locus of altered functional activity in both acute and chronic AN-r, our findings strongly suggest that altered activity in reward-based regions is a consequence of structural changes (namely brain atrophy and alterations in functional connectivity) related to acute undernutrition which take significant time to normalise. Accordingly, residual activity changes can be observed in recovered individuals which may continue to influence cognition for several years. Reward-based abnormalities should therefore be considered as a state marker reflecting illness progression and symptomatic changes, rather than a trait marker of AN-r.

### 4.5. Future directions

Our meta-regression analyses strongly indicate that clinical characteristics are not predictors of differences in true effect sizes. However, it must be noted that only nine of the included studies reported AAO, and only two of these (Scaife et al., 2016; Via et al., 2015) had a mean AAO above 16 years. This is important as the literature suggests the existence of an early onset (EO) group with a mean AAO of 16.2 years (SD = 2.6), and a late onset (LO) group with a mean AAO of 23.6 years (SD = 7.8) (Volpe et al., 2016). Since our analysis did not adequately capture the approximately 25% of individuals with AN-r who can be classified as LO (Volpe et al., 2016), AAO could still be a predictor of effect size. This has important implications in terms of understanding the neurobiology of AN-r, since EO and LO AN-r may be associated with distinct functional alterations. For example, it is possible that whilst those in the EO group display alterations in reward-related neural activity more consistent with reward-contamination theory (Keating, 2010), the LO group may display functional activity more in keeping with a model of anhedonia (Harrison et al., 2014; Wagner et al., 2007). Nonetheless, and importantly in terms of developing new treatments for AN-r, the exclusion of medicated patients or those with psychiatric comorbidities and their relative controls reliably produced the same results as obtained for the entire sample. This indicates that (i) findings of reward-related functional alterations are robust and specifically associated with AN-r and (ii) currently available psychotropic medications are ineffective in reversing reward contamination in AN-r patients.

Whilst attempts to describe a phenomenon as complex as reward processing are necessarily reductionist, what these findings make clear is that improved treatment prospects for the many patients with AN-r who respond only partially, or not at all, to current treatments requires the investigation of pharmacological agents that target different mechanisms than currently available pharmacotherapies. SSRIs may aid weight restoration (Frank, 2020) and through this exert influence on functional alterations in reward-related brain areas associated with anorectic undernutrition. However, they have no *direct* effect on psychological symptoms associated with reward contamination, which may fuel starvation dependence and relapse into the disorder (Gizewski et al., 2010). To promote long-term recovery, it is likely that psychoactive agents that can exert a more direct influence on reward processing, and which offer a truly ‘disease modifying’ mechanism of action rather than simply targeting overt, physical symptoms, will be required.

A key first step towards this goal will likely be clarifying the precise role of dopaminergic signalling in the pathophysiology of AN-r (Keating et al., 2012; Sodersten et al., 2016), with the possibility that dopaminergic agents could then be used to reverse reward contamination in AN-r and support psychological recovery (Frank, 2014).

## 4. CONCLUSION

To our knowledge, this is the first meta-analysis examining reward-processing in restrictive anorexia nervosa as a function of illness stage. We suggest that rather than being a trait marker of AN-r, reward dysfunction is instead a state marker of acute undernutrition which normalises following weight restoration. Nonetheless, it remains unclear whether alterations are premorbid (and then normalise with treatment) or are a consequence of the disorder and associated brain atrophy (McCormick et al., 2008). Furthermore, our results are primarily limited by inconsistencies in the definition of recovery and challenges in determining illness stage. When studying AN, it is inherently difficult to reliably quantify the duration of illness, since diagnosis and treatment initiation may occur several months (or even years) after initial difficulties emerge (Weigel et al., 2014). Additionally, illness course may be interspersed with periods of partial recovery and relapse. Thus, even when clear definitions of ‘recovery’ are provided, these criteria are often based on physical characteristics such as maintenance of a healthy BMI, and do not capture the psychological correlates of recovery.

Despite this conceptual limitation, our findings strongly indicate that altered reward-processing, potentially related to dopamine signalling (Keating et al., 2012), may be a contributing factor in AN-r. If left untreated, these dysfunctional reward signals may contribute to reward contamination (Keating, 2010) and the maintenance of starvation dependence (Fladung et al., 2013). This discovery may allow for the development of tailored pharmacological treatments for AN-r which work in part by reversing alterations to the way in which stimulus valence is assigned.

## Data Availability

The data used in this review are publicly available and specific references to the datasets and data sources used are provided in the references section of the manuscript.

https://doi.org/10.17605/OSF.IO/4JVZA

## Contributions

**Charlotte S. Rye:** Conceptualisation, Methodology, Investigation, Formal Analysis, Visualisation, Writing – Original Draft. **Felippe E. Amorim:** Methodology, Investigation, Writing – Review & Editing. **Laetitia H.E. Ward:** Methodology, Investigation, Writing – Review & Editing. **Amy L. Milton:** Supervision, Writing – Review & Editing.

## Footnotes

1 Here, AN-r and AN-bp are used to refer to the restrictive and binge-purge subtypes, while AN is used to refer to both subtypes.

## References

American Psychiatric Association. (1994). Diagnostic and Statistical Manual of Mental Disorders (4th ed.). American Psychiatric Publishing.

American Psychiatric Association. (2013). DSM-5: Diagnostic and Statistical Manual of Mental Disorders (5th ed.). American Psychiatric Publishing.

Banfield, S. S., & McCabe, M. (2002). An evaluation of the construct of body image. Adolescence, 37(146), 373–393.

Bär, K.-J., de la Cruz, F., Berger, S., Schultz, C. C., & Wagner, G. (2015). Structural and functional differences in the cingulate cortex relate to disease severity in anorexia nervosa. Journal of Psychiatry and Neuroscience, 40(4), 269–279.

Beck, A., Schlagenhauf, F., Wustenberg, T., Hein, J., Kienast, T., & Kahnt, T. (2006). Ventral striatal activation during reward anticipation correlates with impulsivity in alcoholics. Biological Psychiatry, 66, 734–742.

Bergh, C., & Sodersten, P. (1996). Anorexia nervosa, self-starvation and the reward of stress. Nature Medicine, 2(1), 21–22.

Berridge, K. C. (2009). “Liking” and “wanting” food rewards: Brain substrates and roles in eating disorders. Physiology and Behavior, 97(5), 537–550. 10.1016/j.physbeh.2009.02.044

Berridge, K. C., & Kringelbach, M. L. (2015). Pleasure systems in the brain. Neuron, 86(3), 646–664.

Berridge, K. C., Venier, I. L., & Robinson, T. E. (1989). Taste reactivity analysis of 6-hydroxydopamine-induced aphagia: Implications for arousal and anhedonia hypotheses of dopamine function. Behavioural Neuroscience, 103(1), 36–45.

Bischoff-Grethe, A., McCurdy, D., Grenesko-Stevens, E., (Zoe) Irvine, L. E., Wagner, A., Wendy Yau, W. Y., Fennema-Notestine, C., Wierenga, C. E., Fudge, J. L., Delgado, M. R., & Kaye, W. H. (2013). Altered brain response to reward and punishment in adolescents with Anorexia nervosa. Psychiatry Research - Neuroimaging, 214(3), 331–340. 10.1016/j.pscychresns.2013.07.004

Boehm, I., Finke, B., Tam, F. I., Fittig, E., Scholz, M., Gantchev, K., Roessner, V., & Ehrlich, S. (2016). Effects of perceptual body image distortion and early weight gain on long-term outcome of adolescent anorexia nervosa. European Child and Adolescent Psychiatry, 25, 1319–1326.

Broomfield, C., Stedal, K., Touyz, S., & Rhodes, P. (2017). Labeling and defining severe and enduring anorexia nervosa: A systematic review and critical analysis. International Journal of Eating Disorders, 50(6), 611–623. 10.1002/eat.22715

Bubb, E. J., Metzler-Baddeley, C., & Aggleton, J. P. (2018). The cingulum bundle: Anatomy, function, and dysfunction. Neuroscience and Biobehavioral Reviews, 92, 104–27. https://doi-org.ezp.lib.cam.ac.uk/10.1016/j.neubiorev.2018.05.008

Cao, X., Cao, Q., Long, X., Sun, L., Sui, M., Zhu, C., Zuo, X., Zang, Y., & Wang, Y. (2009). Abnormal resting-state functional connectivity patterns of the putamen in medication-naive children with attention deficit hyperactivity disorder. Brain Research, 1303, 195–206. https://doi-org.ezp.lib.cam.ac.uk/10.1016/j.brainres.2009.08.029

Cornelissen, P. L., Johns, A., & Tovée, M. J. (2013). Body size over-estimation in women with anorexia nervosa is not qualitatively different from female controls. Body Image, 10(1), 103–111.

Cowdrey, F. A., Park, R. J., Harmer, C. J., & McCabe, C. (2011). Increased neural processing of rewarding and aversive food stimuli in recovered anorexia nervosa. Biological Psychiatry, 70(8), 736–743. https://doi-org.ezp.lib.cam.ac.uk/10.1016/j.biopsych.2011.05.028

DeLong, M. R. (1973). Putamen: Activity of single units during slow and rapid arm movements. Science, 179(4079), 1240–1242. 10.1126/science.179.4079.1240

Diaz-Marsa, M., Carrasco, J. L., & Saiz, J. (2000). A study of temperament and personality in anorexia and bulimia nervosa. Journal of Personality Disorders, 14, 352–359.

Dillon, D. G., Rosso, I. M., Pechtel, P., Killgore, W. D., Rauch, S. L., & Pizzagalli, D. A. (2014). Peril and pleasure: An RDOC-inspired examination of threat responses and reward processing in anxiety and depression. Depression and anxiety, 31(3), 233–249.

Drewnowski, A., Halmi, K. A., Pierce, B., Gibbs, J., & Smith, G. P. (1987). Taste and eating disorders. American Journal of Clinical Nutrition, 46(3), 442–450.

Egger, M., Smith, G. D., & Phillips, A. N. (1997). Meta-analysis: principles and procedures. BMJ, 315(7121), 1533–1537.

Egger, M., Smith, G. D., Schneider, M., & Minder, C. (1997). Bias in meta-analysis detected by a simple, graphical test. BMJ, 315(7109), 629–634.

Ehrlich, S., Geisler, D., Ritschel, F., King, J. A., Seidel, M., Boehm, I., Breier, M., Clas, S., Weiss, J., Marxen, M., Smolka, M. N., Roessner, V., & Kroemer, N. B. (2015). Elevated cognitive control over reward processing in recovered female patients with anorexia nervosa. Journal of Psychiatry and Neuroscience, 40(5), 307–315. 10.1503/jpn.140249

Espel-Huynh, H. M., Zhang, F., Boswell, J. F., Thomas, J. G., Thompson-Brenner, H., Juarascio, A. S., & Lowe, M. R. (2020). Latent trajectories of eating disorder treatment response among female patients in residential care. The International journal of eating disorders, 53(10), 1647–1656. 10.1002/eat.23369

Evans, M. M., Kim, J., Abel, T., Nickl-Jockschat, T., & Stevens, H. E. (2024). Developmental disruptions of the dorsal striatum in autism spectrum disorder. Biological Psychiatry, 95(2), 102–111.

Everitt, B. J., Cardinal, R. N., Hall, J., Parkinson, J. A., & Robbins, T. W. (2000). Differential involvement of amygdala subsystems in appetitive conditioning and drug addiction. The amygdala: A functional analysis, 353–390.

Fladung, A. K., Schulze, U. M. E., Schöll, F., Bauer, K., & Grön, G. (2013). Role of the ventral striatum in developing anorexia nervosa. Translational Psychiatry, 3(September), 2–7. 10.1038/tp.2013.88

Frank, G. K. W. (2013). Altered brain reward circuits in eating disorders: chicken or egg? Current Psychiatry Report, 15, 396.

Frank, G. K. W. (2014). Could dopamine agonists aid in drug development or anorexia nervosa? Frontiers in Nutrition, 1, 19. 10.3389/fnut.2014.00019

Frank, G. K. W. (2020). Pharmacotherapeutic strategies for the treatment of anorexia nervosa – too much for one drug? Expert Opinion on Pharmacotherapy, 21(9), 1045–1058.

Frank, G. K. W., Bailer, U. F., Henry, S. E., Drevets, W., Meltzer, C. C., Price, J. C., Mathis, C. A., Wagner, A., Hoge, J., Ziolko, S., & Barbarich-Marsteller, N. (2005). Increased dopamine D2/D3 receptor binding after recovery from anorexia nervosa measured by positron emission tomography and [11c] raclopride. Biological Psychiatry, 58(11), 908–912.

Frank, G. K. W., Collier, S., Shott, M. E., & O’Reilly, R. C. (2016). Prediction error and somatosensory insula activation in women recovered from anorexia nervosa. Journal of Psychiatry and Neuroscience, 41(5), 304–311. 10.1503/jpn.150103

Frank, G. K. W., Reynolds, J. R., Shott, M. E., Jappe, L., Yang, T. T., Tregellas, J. R., & O’Reilly, R. C. (2012). Anorexia nervosa and obesity are associated with opposite brain reward response. Neuropsychopharmacology, 37(9), 2031–2046. 10.1038/npp.2012.51

Frank, G.K.W., Shott, M.E., Keffler, C. and Cornier, M.-A. (2016), Extremes of eating are associated with reduced neural taste discrimination. Int. J. Eat. Disord., 49: 603–612. 10.1002/eat.22538

Fuhrmann, D., Knoll, L. J., & Blakemore, S. J. (2015). Adolescence as a sensitive period of brain development. Trends in Cognitive Sciences, 19(10), 558–566.

Gaudio, S., Piervincenzi, C., Beomonte Zobel, B., Romana Montecchi, F., Riva, G., Carducci, F., & Cosimo Quattrocchi, C. (2015). Altered resting state functional connectivity of anterior cingulate cortex in drug naïve adolescents at the earliest stages of anorexia nervosa. Scientific Reports, 5(1), 10818.

Geisler, D., Ritschel, F., King, J. A., Bernardoni, F., Seidel, M., Boehm, I., Runge, F., Goschke, T., Roessner, V., Smolka, M. N., & Ehrlich, S. (2017). Increased anterior cingulate cortex response precedes behavioural adaptation in anorexia nervosa. Scientific Reports, 7(December 2016), 1–10. 10.1038/srep42066

Gillan, C. M. (2021). Recent developments in the habit hypothesis of OCD and compulsive disorders. In N. A. Fineberg & T. W. Robbins (Eds.), The neurobiology and treatment of OCD: Accelerating progress (pp. 147–167). Springer Nature Switzerland AG. 10.1007/7854_2020_199

Gizewski, E. R., Rosenberger, C., de Greiff, A., Moll, A., Senf, W., Wanke, I., Forsting, M., & Herpertz, S. (2010). Influence of satiety and subjective valence rating on cerebral activation patterns in response to visual stimulation with high-calorie stimuli among restrictive anorectic and control women. Neuropsychobiology, 62, 182–192.

Green, M. W., Elliman, N. A., Wakeling, A., & Rogers, P. J. (1996). Cognitive functioning, weight change and therapy in anorexia nervosa. Journal of Psychiatric Research, 30(5), 401–410.

Haddaway, N. R., Page, M. J., Pritchard, C. C., & McGuinness, L. A. (2022). PRISMA2020: An R package and Shiny app for producing PRISMA 2020-compliant flow diagrams, with interactivity for optimised digital transparency and Open Synthesis Campbell Systematic Reviews, 18, e1230. 10.1002/cl2.1230

Hadland, K. A., Rushworth, M. F. S., Gaffan, D., & Passingham, R. E. (2003). The anterior cingulate and reward-guided selection of actions. Journal of Neurophysiology, 89(2), 1161–1164.

Harrison, A., Mountford, V. A., & Tchanturia, K. (2014). Social anhedonia and work and social functioning in the acute and recovered phases of eating disorders. Psychiatry Research, 218, 187–194.

He, J., Li, X., Li, K., Yang, H., & Wang, X. (2024). Abnormal functional connectivity of the putamen in obsessive-compulsive disorder. Journal of Psychiatric Research, 177, 338–345. https://doi-org.ezp.lib.cam.ac.uk/10.1016/j.jpsychires.2024.07.031

Herzog, D. B., Keller, M. B., Sacks, N. R., Yeh, C. J., & Lavori, P. W. (1992). Psychiatric comorbidity in treatment-seeking anorexics and bulimics. Journal of the American Academy of Child and Adolescent Psychiatry, 31, 810–818.

Holsen, L. M., Lawson, E. A., Blum, J., Ko, E., Makris, N., Fazeli, P. K., Klibanski, A., & Goldstein, J. M. (2012). Food motivation circuitry hypoactivation related to hedonic and nonhedonic aspects of hunger and satiety in women with active anorexia nervosa and weight-restored women with anorexia nervosa. Journal of Psychiatry and Neuroscience, 37(5), 322–332.

Jiang, T., Soussignan, R., Carrier, E., & Royet, J.-P. (2019). Dysfunction of the mesolimbic circuit to food odors in women with anorexia and bulimia nervosa: A fMRI study. Frontiers in Human Neuroscience, 13, 117. 10.3389/fnhum.2019.00117

Kaye, W. H., Ebert, M. H., Raleigh, M., & Lake, R. (1984). Abnormalities in CNS monoamine metabolism in anorexia nervosa. Archives of General Psychiatry, 41(4), 350–355. doi:10.1001/archpsyc.1984.01790150040007

Kaye, W. H., Fudge, J. L., & Paulus, M. (2009). New insights into symptoms and neurocircuit function of anorexia nervosa. Nature Reviews Neuroscience, 10(8), 573–584.

Keating, C. (2010). Theoretical perspective on anorexia nervosa: The conflict of reward. In Neuroscience and Biobehavioral Reviews (Vol. 34, Issue 1, pp. 73–79). 10.1016/j.neubiorev.2009.07.004

Keating, C., Tilbrook, Alan. J., Rossell, Susan. L., Enticott, Peter. G., & Fitzgerald, Paul. B. (2012). Reward processing in anorexia nervosa. Neuropsychologia, 50(5), 567–575.

Keel, P. K., Dorer, D. J., Franko, D. L., Jackson, S. C., & Herzog, D. B. (2005). Postremission predictors of relapse in women with eating disorders. American Journal of Psychiatry, 162(12), 2263–2268.

Kennerley, S., Walton, M. E., Behrens, T. E. J., Buckley, M., & Rushworth, M. F. S. (2006). Optimal decision making and the anterior cingulate cortex. Nature Neuroscience, 9(7), 940–947.

King, J. A., Bernardoni, F., Geisler, D., Ritschel, F., Doose, A., Pauligk, S., Pásztor, K., Weidner, K., Roessner, V., Smolka, M. N., & Ehrlich, S. (2020). Intact value-based decision-making during intertemporal choice in women with remitted anorexia nervosa? An FMRI study. Journal of Psychiatry and Neuroscience, 45(2), 108–116. 10.1503/jpn.180252

Kinoshita, M., Nakajima, R., Shinohara, H., Miyashita, K., Tanaka, S., Okita, H., Nakada, M., & Hayashi, Y. (2016). Chronic spatial working memory deficit associated with the superior longitudinal fasciculus: a study using voxel-based lesion-symptom mapping and intraoperative direct stimulation in right prefrontal glioma surgery. Journal of Neurosurgery, 125(4), 1024–1032.

Koh, M. T., Wilkins, E. E., & Bernstein, I. L. (2003). Novel tastes elevate c-fos expression in the central amygdala and insular cortex: implication for taste aversion learning. Behavioral Neuroscience, 117(6), 1416–1422. https://psycnet.apa.org/doi/10.1037/0735-7044.117.6.1416

Lefebvre, A., Traut, N., Pedoux, A., Maruani, A., Beggiato, A., Elmaleh, M., Germanaud, D., Amestoy, A., Ly-Le Moal, M., Chatham, C., Murtagh, L., Bouvard, M., Alisson, M., Leboyer, M., Bourgeron, T., Toro, R., Dumas, G., Moreau, C., & Delorme, R. (2023). Exploring the multidimensional nature of repetitive and restricted behaviors and interests (RRBI) in autism: neuroanatomical correlates and clinical implications. Molecular Autism, 14(45). 10.1186/s13229-023-00576-z

Lozano-Serra, E., Andrés-Perpiña, S., Lázaro-García, L., & Castro-Fornieles, J. (2014). Adolescent anorexia nervosa: cognitive performance after weight recovery. Journal of Psychosomatic Research, 76(1), 6–11.

Luo, X., Mao, Q., Shi, J., Wang, X., & Li, C. R. (2019). Putamen gray matter volumes in neuropsychiatric and neurodegenerative disorders. World Journal of Psychiatry and Mental Health Research, 3(1), 1020.

Lutz, B., Marsicano, G., Maldonado, R., & Hillard, C. J. (2015). The endocannabinoid system in guarding against fear, anxiety and stress. Nature Reviews Neuroscience 2015 16:12, 16(12), 705–718. 10.1038/nrn4036

Madhavan, K. M., McQueeny, T., Howe, S. R., Shear, P., & Szaflarski, J. (2014). Superior longitudinal fasciculus and language functioning in healthy aging. Brain Research, 1562, 11–22. https://doi-org.ezp.lib.cam.ac.uk/10.1016/j.brainres.2014.03.012

McCabe, C., Mishor, Z., Cowen, P. J., & Harmer, C. J. (2010). Diminished neural processing of aversive and rewarding stimuli during selective serotonin reuptake inhibitor treatment. Biological psychiatry, 67(5), 439–445. 10.1016/j.biopsych.2009.11.001

McCormick, L. M., Keel, P. K., Brumm, M. C., Bowers, W., Swayze, V., Andersen, A., & Andreasen, N. (2008). Implications of starvation-induced change in right dorsal anterior cingulate volume in anorexia nervosa. International Journal of Eating Disorders, 41, 602–610.

McFadden, K. L., Tregellas, J. R., Shott, M. E., & Frank, G. K. W. (2014). Reduced salience and default mode network activity in women with anorexia nervosa. Journal of Psychiatry and Neuroscience, 39(3), 179–188.

Mitelman, S. A., Canfield, E. L., Chu, K.-W., Brickman, A. M., Shihabuddin, L., Hazlett, E. A., & Buchsbaum, M. S. (2009). Poor outcome in chronic schizophrenia is associated with progressive loss of volume of the putamen. Schizophrenia Research, 113(2–3), 241–245. https://doi-org.ezp.lib.cam.ac.uk/10.1016/j.schres.2009.06.022

Morgan, H. G., & Russell, G. F. M. (1975). Value of family background and clinical features as predictors of long-term outcome in anorexia nervosa: four-year follow-up study of 41 patients. Psychological Medicine, 5(4), 355–371.

Murao, E., Sugihara, G., Isobe, M., Noda, T., Kawabata, M., Matsukawa, N., Takahashi, H., Murai, T., & Noma, S. (2017). Differences in neural responses to reward and punishment processing between anorexia nervosa subtypes: An fMRI study. Psychiatry and Clinical Neurosciences, 71(9), 647–658. 10.1111/pcn.12537

Naidich, T. P., Krayenbühl, N., Kollias, S., Bou-Haidar, P., Bluestone, A. Y., & Carpenter, D. M. (2013). White matter. In T. P. Naidich, S. Cha, M. Castillo, & J. G. Smirniotopoulos (Eds.), Imaging of the Brain (Expert rad, pp. 205–244). Elsevier Health Sciences. https://doi-org.ezp.lib.cam.ac.uk/10.1016/B978-1-4160-5009-4.50020-0

Nord, C. L., Lawson, R. P., & Dalgleish, T. (2021). Disrupted Dorsal Mid-Insula Activation During Interoception Across Psychiatric Disorders. American Journal of Psychiatry, 178(8), 761–770. 10.1176/appi.ajp.2020.20091340

Nunn, K., Frampton, I. J., Gordon, I., & Lask, B. (2008). The fault is not in her parents but in her insula - A neurobiological hypothesis of anorexia nervosa. European Eating Disorders Review, 16(5), 355–360. 10.1002/erv.890

O’Hara, C. B., Keyes, A., Renwick, B., Leyton, M., Campbell, I. C., & Schmidt, U. (2016). The effects of acute dopamine precursor depletion on the reinforcing value of exercise in anorexia nervosa. PLoS ONE, 11(1), e0145894.

Oldershaw, A., Treasure, J., Hambrook, D., Tchanturia, K., & Schmidt, U. (2011). Is anorexia nervosa a version of autism spectrum disorders? European Eating Disorders Review, 19(6), 462–474. 10.1002/erv.1069

Page, M. J., McKenzie, J. E., Bossuyt, P. M., Boutron, I., Hoffmann, T. C., Mulrow, C. D., Shamseer, L., Tetzlass, J. M., Akl, E. A., Brennan, S. E., & Chou, R. (2021). The PRISMA 2020 statement: an updated guideline for reporting systematic reviews. BMJ, 372. 10.1136/bmj.n71

Paton, J. J., Belova, M. A., Morrison, S. E., & Salzman, C. D. (2006). The primate amygdala represents the positive and negative value of visual stimuli during learning. Nature, 439(7078), 865–870.

Pochon, J. B., Riis, J., Sanfey, A. G., Nystrom, L. E., & Cohen, J. D. (2008). Functional imaging of decision conflict. The Journal of Neuroscience, 28(13), 3468–3473.

Radua, J., Mataix-Cols, D., Phillips, M. L., El-Hage, W., Kronhaus, D. M., Cardoner, N., & Surguladze, S. (2012). A new meta-analytic method for neuroimaging studies that combines reported peak coordinates and statistical parametric maps. European Psychiatry, 27(8), 605–611.

Radua, J., Rubia, K., Canales-Rodriguez, E. J., Pomarol-Clotet, E., Fusar-Poli, P., & Mataix-Cols, D. (2014). Anisotropic kernels for coordinate-based meta-analyses of neuroimaging studies. Frontiers in Psychiatry, 5(13).

Ritschel, F., Geisler, D., King, J. A., Bernardoni, F., Seidel, M., Boehm, I., Vettermann, R., Biemann, R., Roessner, V., Smolka, M. N., & Ehrlich, S. (2017). Neural correlates of altered feedback learning in women recovered from anorexia nervosa. Scientific Reports, 7, 5421. DOI:10.1038/s41598-017-04761-y

Robinson, T. E., & Berridge, K. C. (1993). The neural basis of drug craving: an incentive-sensitization theory of addiction. Brain Research Review, 18, 247–291.

Scaife, J. C., Godier, L. R., Reinecke, A., Harmer, C. J., & Park, R. J. (2016). Differential activation of the frontal pole to high vs low calorie foods: The neural basis of food preference in Anorexia Nervosa? Psychiatry Research, 30(258), 44–53. https://doi-org.ezp.lib.cam.ac.uk/10.1016%2Fj.pscychresns.2016.10.004

Scaife, J., Godier-McBard, L. R., Reinecke, A., Harmer, C., & Park, R. J. (2016). Differentual activation of the frontal pole to high vs low calorie foods: The neural basis of food preference in Anorexia Nervosa? Psychiatry Research: Neuroimaging, 258, 44–53. 10.1016/j.pscychresns.2016.10.004

Scharner, S., & Stengel, A. (2019). Alterations of brain structure and functions in anorexia nervosa. In Clinical Nutrition Experimental (Vol. 28, pp. 22–32). Elsevier Ltd. 10.1016/j.yclnex.2019.02.001

Schmidt, U., & Treasure, J. (2006). Anorexia nervosa: valued and visible. A cognitive-interpersonal maintenance model and its implications for research and practice. British Journal of Clinical Psychology, 45, 343–366.

Seth, A. K., & Critchley, H. D. (2013). Extending predictive processing to the body: emotion as interoceptive inference. Behavioural and Brain Sciences, 36, 227–228.

Shelley, B. P., & Trimble, M. R. (2004). The insular lobe of Reil–its anatamico-functional, behavioural and neuropsychiatric attributes in humans–a review. The World Journal of Biological Psychiatry, 5(4), 176–200. 10.1080/15622970410029933

Sherman, B. J., Savage, C. R., Eddy, K. T., Blais, M. A., Deckersbach, T., Jackson, S. C., Franko, D. L., Rauch, S. L., & Herzog, D. B. (2006). Strategic memory in adults with anorexia nervosa: are there similarities to obsessive compulsive spectrum disorders? International Journal of Eating Disorders, 39(6), 468–476. 10.1002/eat.20300

Simon, J. J., Stopyra, M. A., & Friederich, H.-C. (2019). Neural processing of disorder-related stimuli in patients with anorexia nervosa: A narrative review of brain imagin studies. Journal of Clincal Medicine, 8(7), 1047.

Smith, D. M., & Torregrossa, M. M. (2021). Valence encoding in the amygdala influences motivated behaviour. Behavioural Brain Research, 411, 113370. 10.1016/j.bbr.2021.113370

Smith, K. S., Berridge, K. C., & Aldridge, J. W. (2011). Disentangling pleasure from incentive salience and learning signals in brain reward circuitry. Proceedings of the National Academy of Sciences of the United States of America, 108(27), E255–E264.

Sodersten, P., Bergh, C., Leon, M., & Zandian, M. (2016). Dopamine and anorexia nervosa. Neuroscience and Biobehavioral Reviews, 60, 26–30.

Strigo, I. A., Matthews, S. C., Simmons, A. N., Oberndorfer, T., Klabunde, M., Reinhardt, L. E., & Kaye, W. H. (2013). Altered insula activation during pain anticipation in individuals recovered from anorexia nervosa: Evidence of interoceptive dysregulation. International Journal of Eating Disorders, 46(1), 23–33. 10.1002/eat.22045

Su, T., Gong, J., Tang, G., Qiu, S., Chen, P., Chen, G., Wang, J., Huang, L., & Wang, Y. (2021). Structural and functional brain alterations in anorexia nervosa:A multimodal meta-analysis of neuroimaging studies. In Human Brain Mapping (Vol. 42, Issue 15, pp. 5154–5169). John Wiley and Sons Inc. 10.1002/hbm.25602

Suchan, B., Bauser, D. S., Busch, M., Schulte, D., Gronemeyer, D., Herpertz, S., & Vocks, S. (2013). Reduced connectivity between the left fusiform body area and the extrastriate body area in anorexia nervosa is associated with body image distortion. Behavioural Brain Research, 241, 80–85.

Sweitzer, M. M., Watson, K. K., Erwin, S. R., Winecoff, A. A., Datta, N., Huettel, S., Platt, M. L., & Zucker, N. L. (2018). Neurobiology of social reward valuation in adults with a history of anorexia nervosa. PLoS ONE, 13(12), 1–22. 10.1371/journal.pone.0205085

Tauro, J. L., Wearne, T. A., Belevski, B., Filipčíková, M., & Francis, H. M. (2022). Social cognition in female adults with Anorexia Nervosa: A systematic review. Neuroscience & Biobehavioral Reviews, 132, 197–210. https://doi-org.ezp.lib.cam.ac.uk/10.1016/j.neubiorev.2021.11.035

Treasure, J., & Schmidt, U. (2013). The cognitive-interpersonal maintenance model of anorexia nervosa revisited: a summary of the evidence for cognitive, socio-emotional and interpersonal predisposing and perpetuating factors. Journal of Eating Disorders, 1, 1–10.

Van Autreve, S., De Baene, W., Baeken, C., van Heeringen, C., & Vervaet, M. (2013). Do restrictive and bingeing/purging subtypes of anorexia nervosa differ on central coherence and set shifting? European Eating Disorder Review, 21, 308–314.

van Eeden, A. E., van Hoeken, D., & Hoek, H. W. (2021). Incidence, prevalence and mortality of anorexia nervosa and bulimia nervosa. Current Opinion in Psychiatry, 34(6), 515–524. 10.1097/YCO.0000000000000739

Via, E., Soriano-Mas, C., Sánchez, I., Forcano, L., Harrison, B. J., Davey, C. G., Pujol, J., Martínez-Zalacaín, I., Menchón, J. M., Fernández-Aranda, F., & Cardoner, N. (2015). Abnormal social reward responses in anorexia nervosa: An fmri study. PLoS ONE, 10(7), 1–20. 10.1371/journal.pone.0133539

Volpe, U., Tortorella, A., Manchia, M., Monteleone, A. M., Albert, U., & Monteleone, P. (2016). Eating disorders: What age at onset? Psychiatry Research, 238, 225–227. https://doi-org.ezp.lib.cam.ac.uk/10.1016/j.psychres.2016.02.048

Wagner, A., Aizenstein, H., Mazurkewicz, L., Fudge, J., Frank, G. K., Putnam, K., Bailer, U. F., Fischer, L., & Kaye, W. H. (2008). Altered insula response to taste stimuli in individuals recovered from restricting-type anorexia nervosa. Neuropsychopharmacology, 33(3), 513–523. 10.1038/sj.npp.1301443

Wagner, A., Aizenstein, H., Venkatraman, V. K., Fudge, J., May, J. C., Mazurkewicz, L., Frank, G. K., Bailer, U. F., Fischer, L., Nguyen, V., Carter, C., Putnam, K., & Kaye, W. H. (2007). Altered reward processing in women recovered from anorexia nervosa. American Journal of Psychiatry, 164(12), 1842–1849. 10.1176/appi.ajp.2007.07040575

Wagner, A., Greer, P., Bailer, >Ursula. F., Frank, G. K., Henry, S. E., Putnam, K., Meltzer, C. C., Zolko, S. K., Hoge, J., McConaha, C., & Kaye, W. K. (2006). Normal brain tissue volumes after long-term recovery in Anorexia and Bulimia Nervosa. Biological Psychiatry, 59(3), 291–293.

Weigel, A., Rossi, M., Wendt, H., Neubauer, K., von Rad, K., Daubmann, A., Romer, G., Löwe, B., & Gumz, A. (2014). Duration of untreated illness and predictors of late treatment initiation in anorexia nervosa. Journal of Public Health (Germany), 22(6), 519–527. 10.1007/s10389-014-0642-7

Whishaw, I. Q., O’Connor, W. T., & Dunnett, S. B. (1986). The contributions of motor cortex, nigrostriatal dopamine and caudate-putamen to skilled forelimb use in the rat. Brain, 109(5), 805–843.

Wilson, D. R., Loxton, N. J., O’Shannessy, D., Sheeran, N., & Morgan, A. (2019). Similarities and differences in revised reinforcement sensitivities across eating disorder subtypes. Appetite, 133, 70–76. 10.1016/j.appet.2018.10.023

Wrase, J., Kahnt, T., Schlagenhauf, F., Beck, A., Cohen, M., Knutson, B., & Heinz, A. (2007). Different neural systems adjust motor behavior in response to reward and punishment. NeuroImage, 36, 1253–1262.

Zhong, S., Su, T., Gong, J., Huang, L., & Wang, Y. (2023). Brain functional alterations in patients with anorexia nervosa: A meta-analysis of task-based functional MRI studies. Psychiatry Research, 327(December 2022), 115358. 10.1016/j.psychres.2023.115358

Zhu, Y., Hu, X., Wang, J., Chen, J., Guo, Q., Li, C., & Enck, P. (2012). Processing of food, body and emotional stimuli in anorexia nervosa: a systematic review and meta-analysis of functional magnetic resonance imaging studies. European Eating Disorders Review, 20(6), 439–450.

Zipfel, S., Giel, K. E., Bulik, C. M., Hay, P., & Schmidt, U. (2015). Anorexia nervosa: Aetiology, assessment and treatment. Lancet Psychiatry, 2, 1099–1111.

