## Supplementary Material 1 for "Reward contamination in restrictive anorexia nervosa: A meta-analysis of functional MRI studies"

### Supplementary Material 1. Quality Assessment Checklist (Zhong et al., 2023): Score 0/0.5/1 per item; total score out of 12

---

**Category 1: Subjects Score**

1. Participants were evaluated prospectively, specific diagnostic criteria were applied (DSM-IV/V), and demographic data was reported.
2. Healthy comparison subjects were evaluated prospectively, psychiatric, and medical illnesses were excluded, and demographic data was reported.
3. Important variables (e.g. clinical scales, illness history, number of episodes; drug status, non-drug therapy status) were checked either by stratification or statistically.
4. Sample size per group > 10, and no significant difference in age and sex existed.

**Category 2: Methods for image acquisition and analysis**

5. Magnet strength at least 1.5T.
6. Whole brain analysis was automated with no a-priori regional selection.
7. Coordinates reported in a standard space (in condition of no significant differences between groups, this item would also be regarded met since no coordinates need to be reported).
8. The imaging technique used was clearly described so as it could be reproduced.
9. Measurements were clearly described so that they could be reproduced.
10. Results have been corrected for multiple comparison

**Category 3: Results and conclusions**

11. Statistical parameters for significant and important non-significant differences were provided.
12. Conclusions were consistent with the results obtained and the limitations were discussed.
